# Reevaluating the Role of ACE Inhibitors in Skin Fibrosis Risk: Evidence from Mendelian Randomization

**DOI:** 10.1101/2024.07.23.24310902

**Authors:** Yangyang Wei, Ziqi Wan, Yiwen Jiang, Zhengye Liu, Ming Yang, Jieying Tang

## Abstract

**Background:** Skin fibrosis, characterized by excessive extracellular matrix deposition, leads to hypertrophic scars and keloids, which are both common and often detrimental conditions. Angiotensin-converting enzyme (ACE) inhibitors have shown promise in animal studies and limited clinical trials for reducing scar formation. However, the causal relationship between ACE inhibition and skin fibrosis remains unclear.

**Methods:** This study employed two-sample Mendelian randomization (MR) analysis to investigate the causal effect of ACE inhibition on skin fibrotic diseases. We utilized genetic variants associated with serum ACE levels, ACE inhibition, and effect of decreasing blood pressure by ACE inhibition as instrumental variables. We analyzed the association between these exposures and the incidence of skin fibrosis, hypertrophic scars, and keloids using various MR methods.

**Results:** We found no significant causal relationship between genetically proxied serum ACE levels, or local skin tissue ACE expression and the risk of skin fibrosis, hypertrophic scars, or keloids. Additionally, there was no direct causal relationship between the effect of ACE inhibitors on blood pressure reduction and the risk of skin fibrotic diseases. However, we observed a significant negative association between systolic blood pressure (SBP) and the risk of hypertrophic scars. Conversely, we found a positive association between β-blockers and the risk of skin fibrosis.

**Conclusion:** Our findings suggest that ACE inhibitors do not have a direct causal effect on the risk of skin fibrotic diseases, including hypertrophic scars and keloids. This challenges the potential of ACE inhibitors as a therapeutic option for preventing or treating these conditions.

## Introduction

Skin fibrosis involves the deposition of extracellular matrix of dermis, a process that can occur in all kinds of wound healing of injuries.^1^ Hypertrophic scars and keloids are two common types of excessive scar caused by skin fibrosis. Hypertrophic scar is that scar proliferation stays within the limits of the edges of the initial scar, whereas keloid is characterized by pseudo-tumor proliferation extending over the edges of the initial wound. ^1, 2^ These conditions are highly prevalent in population, affecting patients with all kinds of skin wounds including surgical ones. Excessive scars can lead to unexpected changes in appearance, causing pain and functionally impairing patients’ quality of life. Current conservative treatments for excessive scarring include cryotherapy, corticosteroid injections or tapes, chemotherapy and surgery. ^1^ Few interventions target the molecular mechanism specifically. Mechanisms of skin fibrosis are not completely understood, involving intricate pathways such as inflammatory reactions and hormone regulation. ^3^

Angiotensin-converting enzyme (ACE) is a well-known molecule that can increase blood pressure by converting angiotensin I to angiotensin II within renin-angiotensin system (RAS). ^4^ Angiotensin II can bind to the Type I angiotensin receptor (AT1 receptor) and initiate the complex downstream effects. The serum ACE has been demonstrated to be critical in the development and progression of fibrosis in multiple organs, including heart, liver, and kidney. ^5–9^ Effects of angiotensin II include the increase of collagen and fibronectin synthesis, which are ultimately associated with fibrosis in different organs. Given its pivotal role in the regulation of RAS, ACE has been inferred to be a therapeutic potential target for skin fibrosis due to its effects on inflammatory reaction and homeostasis of skin healing. ^5^ Animal studies have provided evidence that ACE gene deficiency decreases scar formation in rats effectively. ^11, 12^ A study on rabbits also demonstrated that oral ACE inhibitor (Enalapril) effectively decreased hypertrophic scars on rabbit ears. ^13^ And several clinical researches reported the effectiveness of ACE inhibitors in decreasing the formation of excessive scars. A retrospective chart review study on bilateral breast reduction surgery (n = 961) discovered patients with an ACE inhibitors or angiotensin receptor blocker (ARB) exhibited a significantly lower incidence of being diagnosed as hypertrophic scar. ^14^ Furthermore, in another double-blind clinical trial conducted in Iran population (n = 30), patients treated with local topical Enalapril exhibited hypertrophic scars with significantly smaller mean thickness than the placebo-treated ones, along with a significantly lower mean itching score. ^15^ However, the sample sizes of the clinical trial were insufficiently large and only involved Iran population. The researches were limited by study design. ^16^

Mendelian randomization is a method to explore the causal effect of two traits (exposure and outcome) via genetic instrumental variants (IVs). ^17, 18^ It holds three assumptions: 1) IVs are extensively associated with exposure; 2) There are no confounders between the instruments and outcome; 3) IVs’ effect on outcome can only be exerted via exposure. MR provides a convenient method for exploring the causal relationship between phenotypes with evidence of nearly the same level as randomized controlled trial (RCT) when conducting RCTs is challenging. However, there has not been an exploration of the causal relationship between ACE inhibition and diseases related to skin fibrosis using MR analysis.

Here we aimed to explore the causal effect of ACE inhibitors (ACEi) on excessive scars related to skin fibrosis, including hypertrophic scar and keloids, by two-sample MR analysis from multiple dimensions. We investigated the causal effects of genetically proxied serum ACE, inhibition of ACE and systolic and diastolic blood pressure (SBP/DBP), respectively, on the incidence of skin fibrosis. And we further explored the causal effects of SBP, DBP, β blockers and calcium channel blockers, angiotensin receptor blockers and excessive scarring related to skin fibrosis.

## Methods

### Study design

The study design is illustrated in Figure 1. To investigate the causal effect of ACE inhibitors on skin fibrotic diseases, we conducted MR analysis with exposures of serum ACE, genetically-proxied inhibition of serum ACE, and effect of increasing SBP and DBP by ACE, respectively. Furthermore, we employed Summary-based MR analysis (SMR) to explore the causal effect of ACE expression in local skin tissue on skin fibrotic diseases. Considering that the target of Type 1 angiotensin II receptor blockers (ARBs), AT1 receptor is the main downstream receptor of angiotensin II, we investigated causal relationship between ARBs and skin fibrotic diseases. Moreover, considering that blood pressure might be a risk factor of skin fibrotic diseases ^19, 20^, in order to investigate whether the effect of ACE on skin fibrotic diseases is through the blood pressure, we also investigated the causal effect of blood pressure and targets of other anti-hypertension drugs (β blockers and calcium channel blockers, abbreviated as BBs and CCBs) on skin fibrotic diseases through MR analysis. All the genetic IVs were chosen on autosomes in order to avoid sex-specific effects. ^21^

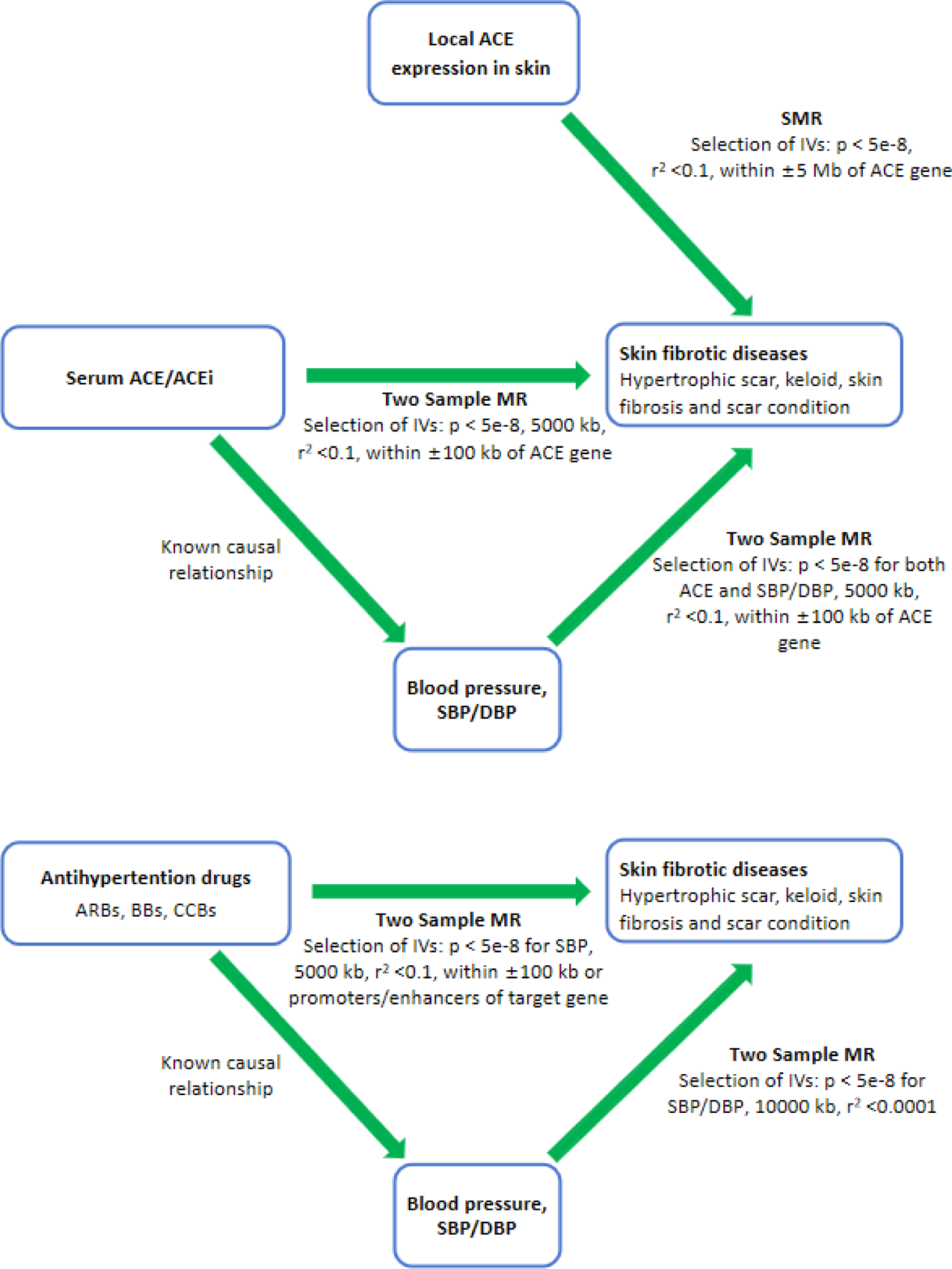

### Data sources

For serum ACE concentration, we initially utilized single SNP “rs4343” as IV of serum ACE. Rs4343 is located in the *ACE* gene, which is extensively associated with an indel known as *Alu* in the 16^th^ intron and then asscociated with expression of *ACE*. ^23^ Rs4343 exhibits the strongest association with serum ACE inhibition (p = 1.53e-213) among SNPs according to the group of IV proxied serum ACE inhibition. ^24^ Additionally, we utilized another group of IVs from a protein quantitative trait locus (pQTL) research of serum ACE of 35,275 individuals with Island ancestry. The population shares a similar age, gender, and BMI structure with the UKB corhort. ^22^ Furethermore, in order to supply more evidence of the causal relationship between serum ACE inhibition and skin fibrotic diseases, we utilized a group of IVs (including rs4343) to proxy serum ACE inhibition from a previous MR study, which was from a pQTL of 4,174 individuals from Genetic Epidemiology Research on Adult Health and Aging. ^24^

We initially employed single SNP “rs4291” to proxy the effect of ACE inhibition on decreasing SBP. Rs4291 is located on the promoter of ACE gene, which is also extensively associated with the indel *Alu* and exhibits the most significant correlation with the effect of ACE inhibition on decreasing SBP. ^25^ Its effect was from a meta-analysis of GWAS covered 757,601 individuals with European ancestry of UKB individuals and International Consortium of Blood Pressure GWAS. ^26^ Then we utilized another group of IV associated with the decrease effect of SBP by ACE (containing 2 SNPs) extracted by a previous MR study, whose effect was from the mentioned meta-analysis covered 757,601 individuals. ^24^ We accessed GWAS data of SBP and DBP from a previous study encompassing 469,767 and 490,469 individuals within European ancestry in UK Biobank cohort. ^28^ And we selected SNPs that have strong relevance (p < 5e-8) with serum ACE according to the mentioned pQTL, and extracted their effects on SBP or DBP from these two GWAS datasets of SBP and DBP. From this set of SNPs, we further included SNPs that also had strong relevance of SBP or DBP (p < 5e-6) to proxy the ACE-induced increase of SBP or DBP.

For local expression of *ACE* gene in skin tissue, summary data of expression quantitative trait locus research (eQTL) of local tissue was downloaded from GTEx v8 data. ^29^ Considering the influence of sunshine exposure on skin, we employed summary data from suprapubic skin and skin of lower leg to represent skin with and without sunshine exposure respectively.

In order to investigate the potential causal effect of anti-hypertension drugs (BBs and CCBs) on skin fibrotic diseases, we extracted IVs proxied the effect of increasing SBP by BB and CCB drug target gene (*ADRB1* for BB, *CACNA1D, CACNA1F, CACNA2D1, CACNA2D2, CACNA1S, CACNB1, CACNB2, CACNB3, CACNB4, CACNG1*, and *CACNA1C* for CCB) from a previous study of MR analysis. ^26^ IV for effect of decreasing SBP by ARBs target gene (type 1 angiotensin II receptor, *AT1*) were extracted from another MR study. ^27^ All the effect data to SBP was from the meta-analysis of GWAS covered 757,601 individuals All the detailed information for exposures were listed in Table 1.

**Table 1.**
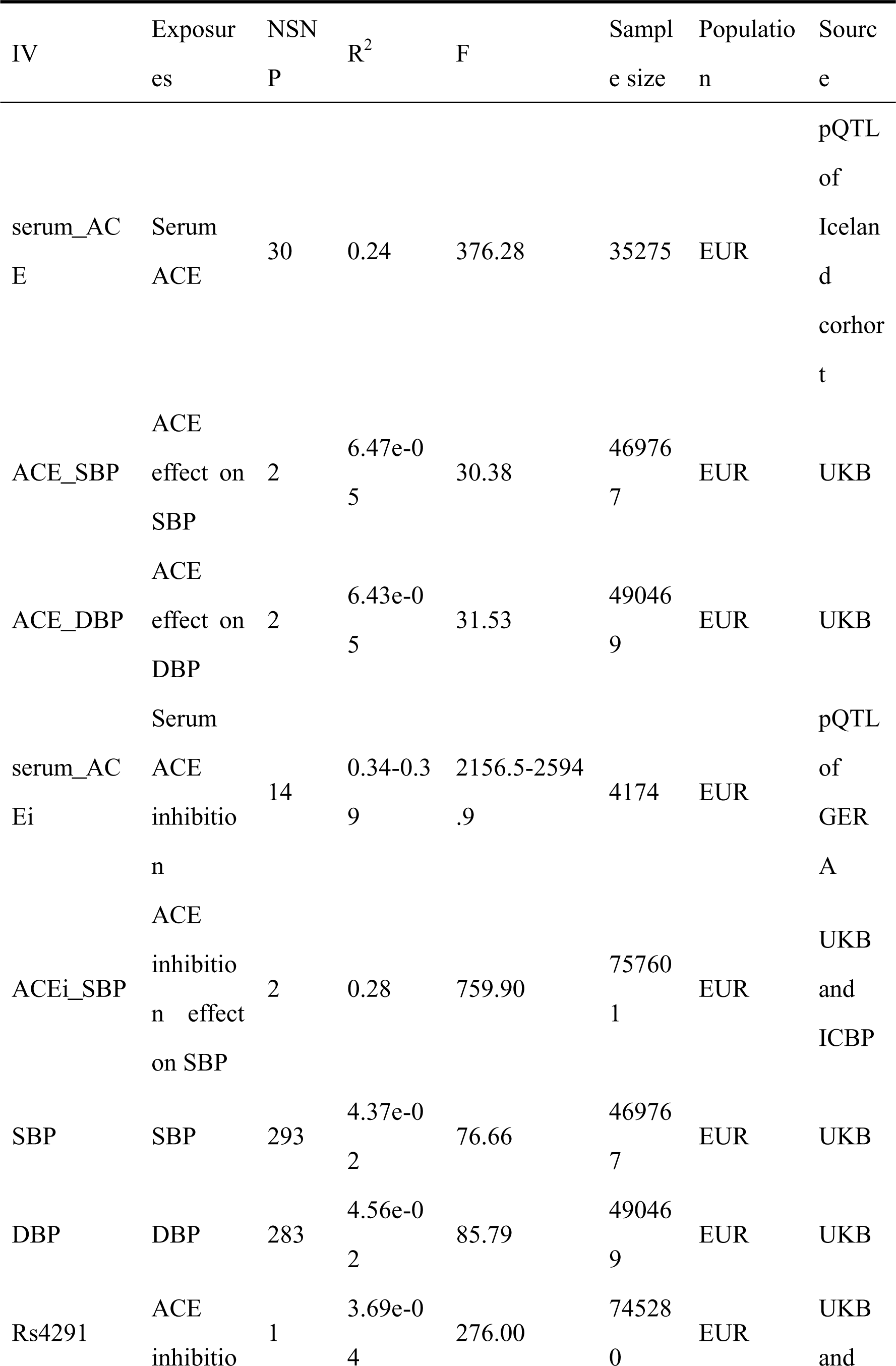
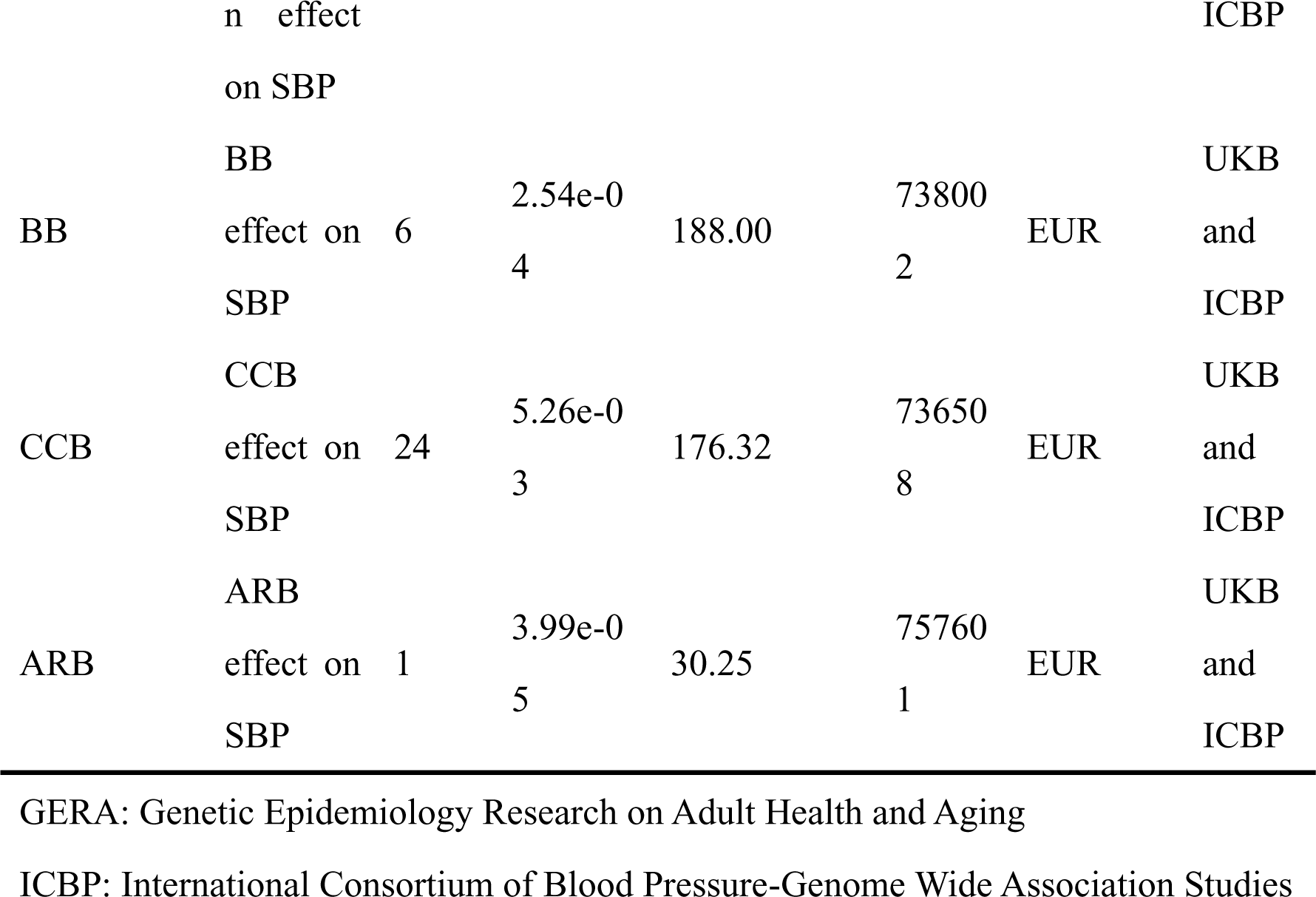
Detailed Information for Exposure GWAS Data.

We employed GWAS datasets for hypertrophic scar, keloid, skin fibrosis, and scar condition (defined as a fibrotic scar that could not be differentiated as keloid or hypertrophic scar). ^1^ All the data were sourced from populations of European ancestry. Among these data for hypertrophic scar and one data of skin fibrosis and scar condition were from FinnGen, whereas data for keloid and the other data of skin fibrosis and scar condition were from UK Biobank. The detailed information for each dataset was listed in Table 2. ^28, 30^

**Table 2.** Detailed Information for Outcome GWAS Data.

| Phenotype | UKB-id | Sample<br>size | Ncase | Ncontrol | Source | Population |
| --- | --- | --- | --- | --- | --- | --- |
| Hypertrophic scar | - | 323,197 | 1294 | 321903 | Finngen | EUR |
| Keloid | 701.4 | 404,486 | 216 | 402139 | UKB | EUR |
| Skin fibrosis and scar<br>condition | 701.2 | 404,486 | 2345 | 402141 | UKB | EUR |
| Skin fibrosis and scar<br>condition | - | 322,794 | 891 | 321903 | Finngen | EUR |

### MR IVs selection

For IVs of multiple SNPs proxied effects of target genes (ACE, target genes for BBs, CCBs and ARBs), we selected SNPs with strong relevance (p < 5e-8) to respective exposures. For serum ACE and serum ACE inhibition, we selected SNPs within ± 100 kb of *ACE* gene. For effects of decreasing SBP by BBs and CCBs, we selected SNPs within corresponding genes or in the promoters and enhancers. ^26^ IV for ARB was selected in the drug-targeted genes associated with BP at a genome-wide significant level, which was identified in DrugBank. ^27^ Promoter and enhancer regions of corresponding drug targets were identified using the GeneHancer database in the GeneCards online platform. Subsequently, we decreased the linkage disequilibrium of the IVs by clumping on condition that genetic distance > 10,000 kb and r^2^ < 0.1.

From the GWAS data of SBP and DBP, we selected SNPs with strong relevance (p < 5e-8). ^31^ Then SNPs was clumped on condition that genetic distance > 10,000 kb and r^2^ < 0.0001 to eliminate linkage disequilibrium.

For IV of effect of serum ACE on decreasing SBP and DBP, we extracted SNPs with extensive relevance for serum ACE (data from pQTL of serum ACE, p < 5e-8) and SBP or DBP (data from GWAS of SBP and DBP, p < 5e-6, for no SNP existed on condition that p < 5e-8). Then we selected SNPs within ± 100 kb of ACE gene and clumped on condition that genetic distance > 10,000 kb and r^2^ < 0.1 thereby reducing linkage disequilibrium.

For all IVs that will be used in MR analysis, we harmonized effects of SNPs on exposure and outcome to ensure effects relative to the same allele and exclude palindromic SNPs with intermediate allele frequencies (> 0.42). In order to avoid potential confounding, we delete SNPs strongly related (p < 5e-8) to outcomes (skin fibrosis, hypertrophic scar and keloid) by investigating all the SNPs on PhenoScanner database (http://www.phenoscaner.medschl.cam.ac.uk/). ^32^ For the exposure of SBP, DBP and the effect on SBP/DBP by ACEi, ARB, BBs and CCBs included UKB cohorts, MR analysis with sample overlapping (outcome data from UKB cohorts) was eliminated by checking the sample sources of exposures and outcomes from different cohorts.

### Statistics

R (Version 4.3.3, The R Foundation for Statistical Computing Platform, Vienna, Austria) was used to perform MR analysis and subsequent sensitivity analysis. R packages “dplyr”, “TwoSampleMR” (Version 0.6.4) and “ieugwasr” ^33^ were used to input, clump and analyze data. To explore the causal relationship of exposures and outcomes, when only one SNP existed in IV, we used method Wald ratio, while when two or more SNPs existed, we used method inverse variance weighted (IVW) including random effects IVW (IVW-MRE) and fixed effects IVW (IVW-FE). ^34^ In order to obtain more reference, we performed MR analysis by the weighted-median ^35^, MR Egger ^36^, simple-mode and weighted-mode method as secondary analysis methods. In MR analysis between drug targets (ACEi, ARBs, BBs and CCBs), we didn’t employ the MR Egger due to the confounding effects of IVs against the INSIDE assumptions of MR Egger. Cochran’s Q statistic was used to estimate the heterogeneity of IVs. If no significant heterogeneity exists (pQ < 0.05), we referred to results from the fixed-effects IVW MR analysis. If significant heterogeneity exists (pQ < 0.05), we referred to results from the random-effects IVW MR analysis. And we tested the pleiotropy of SNPs by the intercept value from the MR Egger method ^36–38^. The p value less than 0.05 indicated significant pleiotropy. If IVs of exposure were with pleiotropy, the analysis results would be excluded. F statistics were used to evaluate the risk of weak instrument bias. We used formula that F = (N-k-1)/k*r^2^/(1-r^2^) to calculate F. Among those N is the sample size of GWAS research, k is the amount of IVs. For each IV, r^2^ was calculated as 2*MAF*(1-MAF)*β^2^. ^39^ If F statistics of a group of IVs was above 10, we considered that risk of weak instrument bias of IV can be ignored. An online tool (https://sb452.shinyapps.io/power/) was used to calculate the statistical power for MR analysis. ^40^

Summary-based MR (SMR) analysis was conducted on SMR software (version 1.3.1 for Windows system) developed by Zhang et al. ^41^ The software extracted SNPs within ± 5 Mb of ACE gene and with significant relevance of ACE expression in specific tissue (p < 5e-8) from the two eQTL datasets from skin tissue. We excluded SNPs whose allele frequency below 0.01. Then we reduced LD (R^2^ < 0.1) by referring to the 1000G European data. GWAS datasets of skin fibrotic diseases were used as outcomes in SMR test. Then we conducted HEIDI test to test the heterogenesis. If pHEIDI was above 0.05, we considered the IVs exhibit no significant heterogenesis. Results of SMR with significant heterogenesis were removed.

## Results

### IVs selection

All the F statistics of IVs were above 10 (listed in Table 1), and the explained variance of IVs were R^2^ of 0.025% to 39%. Detailed information including amounts of SNPs, R^2^, F statistics, sample size and source were shown in Table 1. All the SNPs used in MR were listed in Supplementary File 1. MR analysis whose exposure and outcome were with sample overlapping was removed.

### Effect of serum ACE or ACE inhibition and skin fibrotic diseases

Our analysis of MR results revealed little causal relationship between serum ACE or ACE inhibition and skin fibrotic diseases. For single SNP (rs4343) proxied serum ACE inhibition, results of Wald ratio showed little causal effect on skin fibrotic diseases (hypertrophic scar: OR = 1.04, 95%CI 0.93-1.17, p = 0.46; keloid: OR = 1.12, 95%CI 0.83-1.52, p = 0.45; skin fibrosis and scar condition from UKB: OR = 0.95, 95%CI 0.39-2.35, p = 0.30; skin fibrosis and scar condition from FinnGen: OR = 0.93, 95%CI 0.81-1.07, p = 0.29). For IVs composed of multiple SNPs proxied serum ACE, results of IVW showed little causal effect on skin fibrotic diseases (hypertrophic scar: OR = 0.96, 95%CI 0.86-1.07, p = 0.44; keloid: OR = 1.03, 95%CI 0.78-1.35, p = 0.49; skin fibrosis and scar condition from UKB: OR = 0.98, 95%CI 0.90-1.06, p = 0.56; skin fibrosis and scar condition from FinnGen: OR = 1.11, 95%CI 0.98-1.25, p = 0.12). For IVs containing multiple SNPs proxied serum ACE inhibition, we found little causal effect of ACE inhibition on skin fibrotic diseases (hypertrophic scar: OR = 0.998, 95%CI 0.924-1.078, p = 0.96; keloid: OR = 1.05, 95%CI 0.85-1.29, p = 0.66; skin fibrosis and scar condition from UKB: OR = 0.97, 95%CI 0.91-1.03, p = 0.33; skin fibrosis and scar condition from FinnGen: OR = 0.94, 95%CI 0.85-1.03, p = 0.16). Additionally, other methods also showed little causal relationship between serum ACE or ACE inhibition and skin fibrotic diseases. No significant pleiotropy or heterogeneity bias was observed in these MR analysis (shown in Table 3).

**Table 3.**
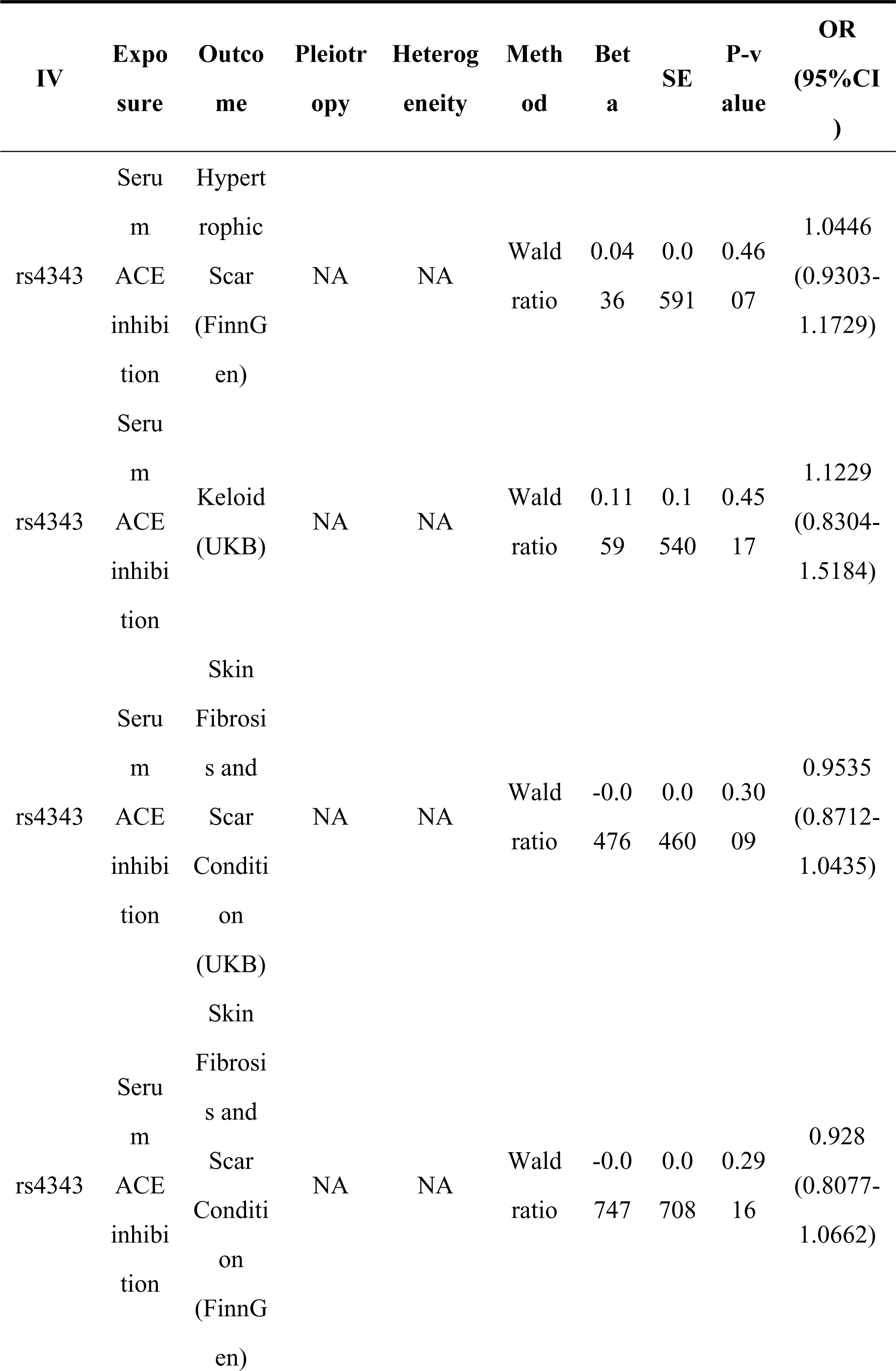
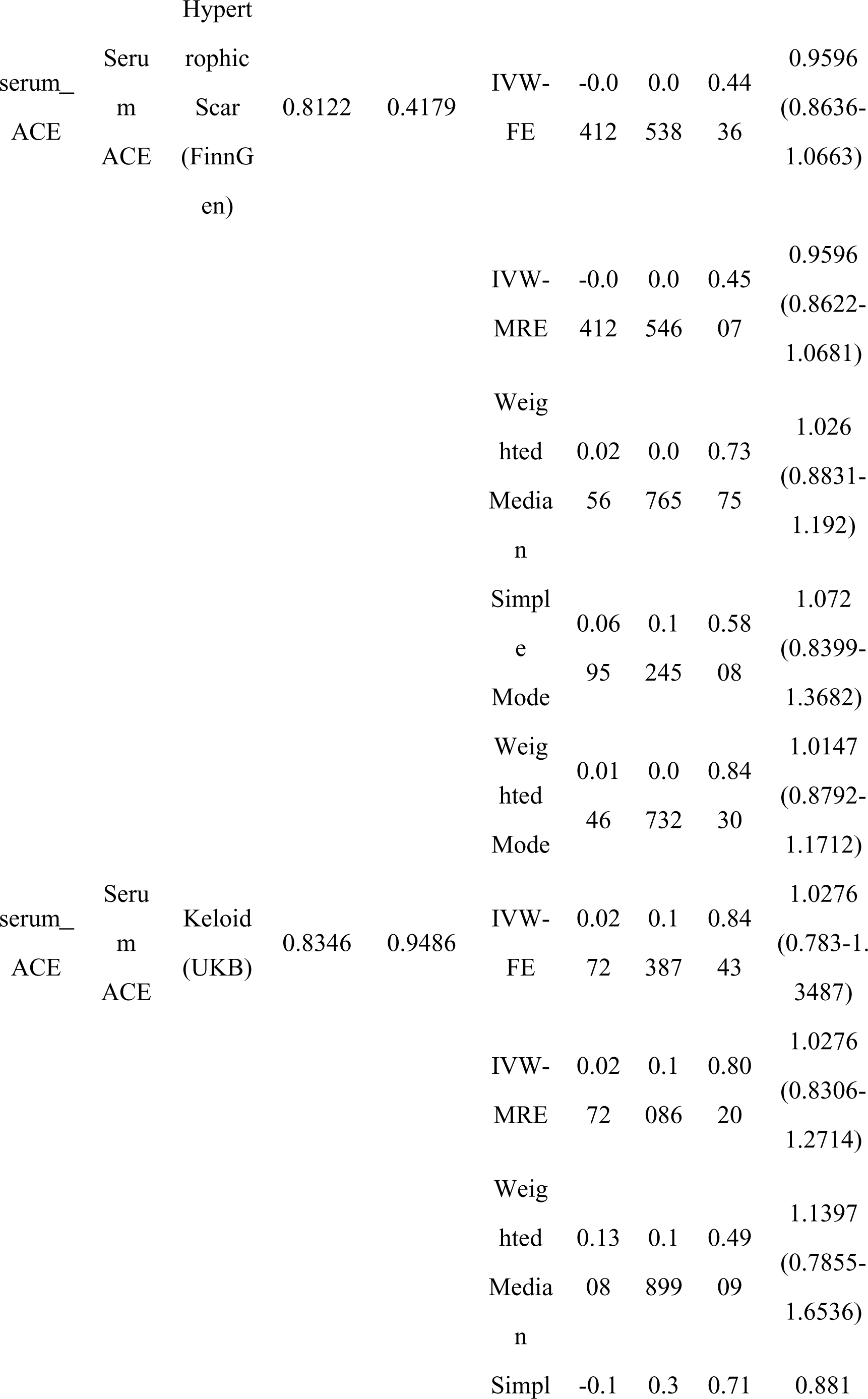
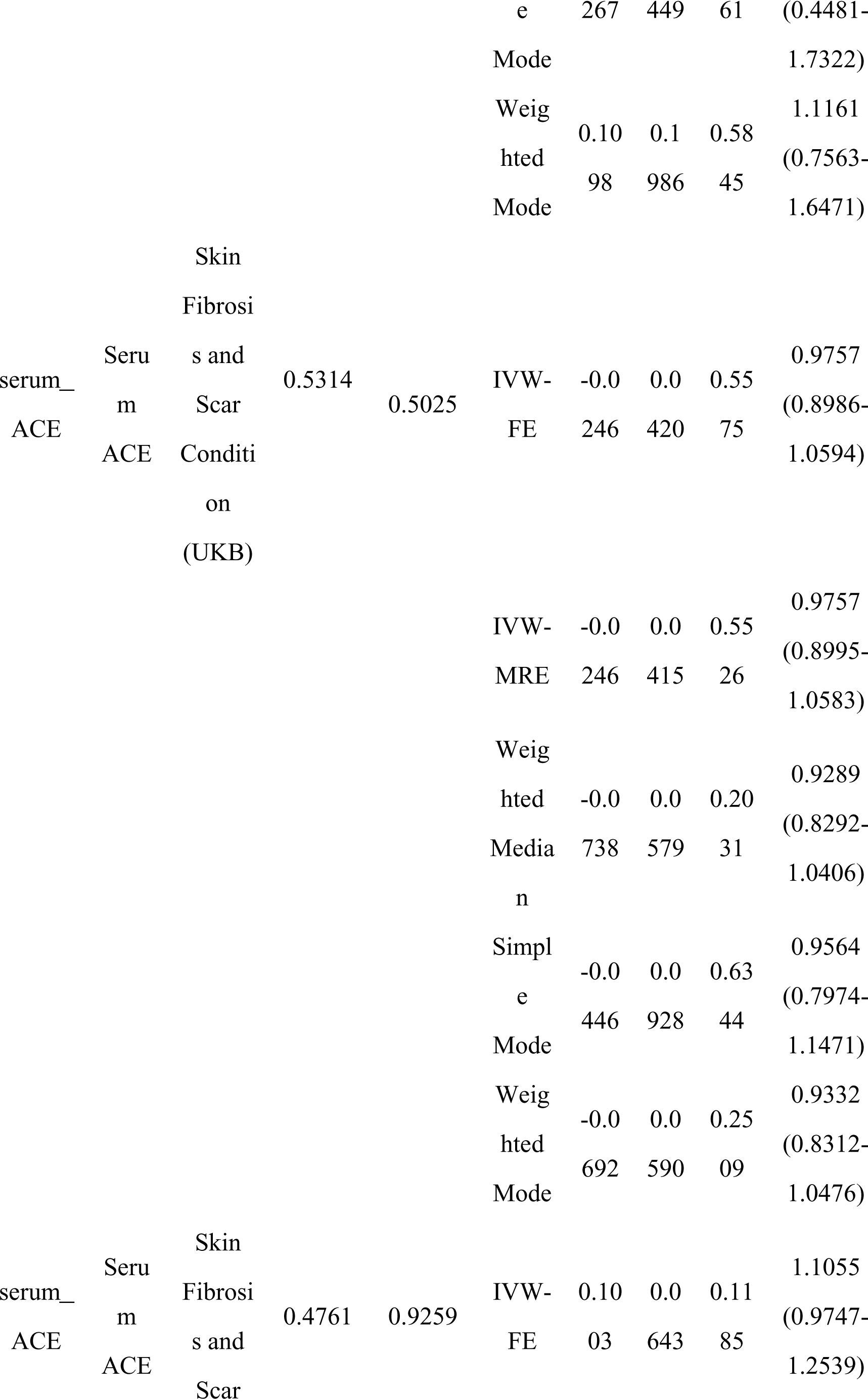
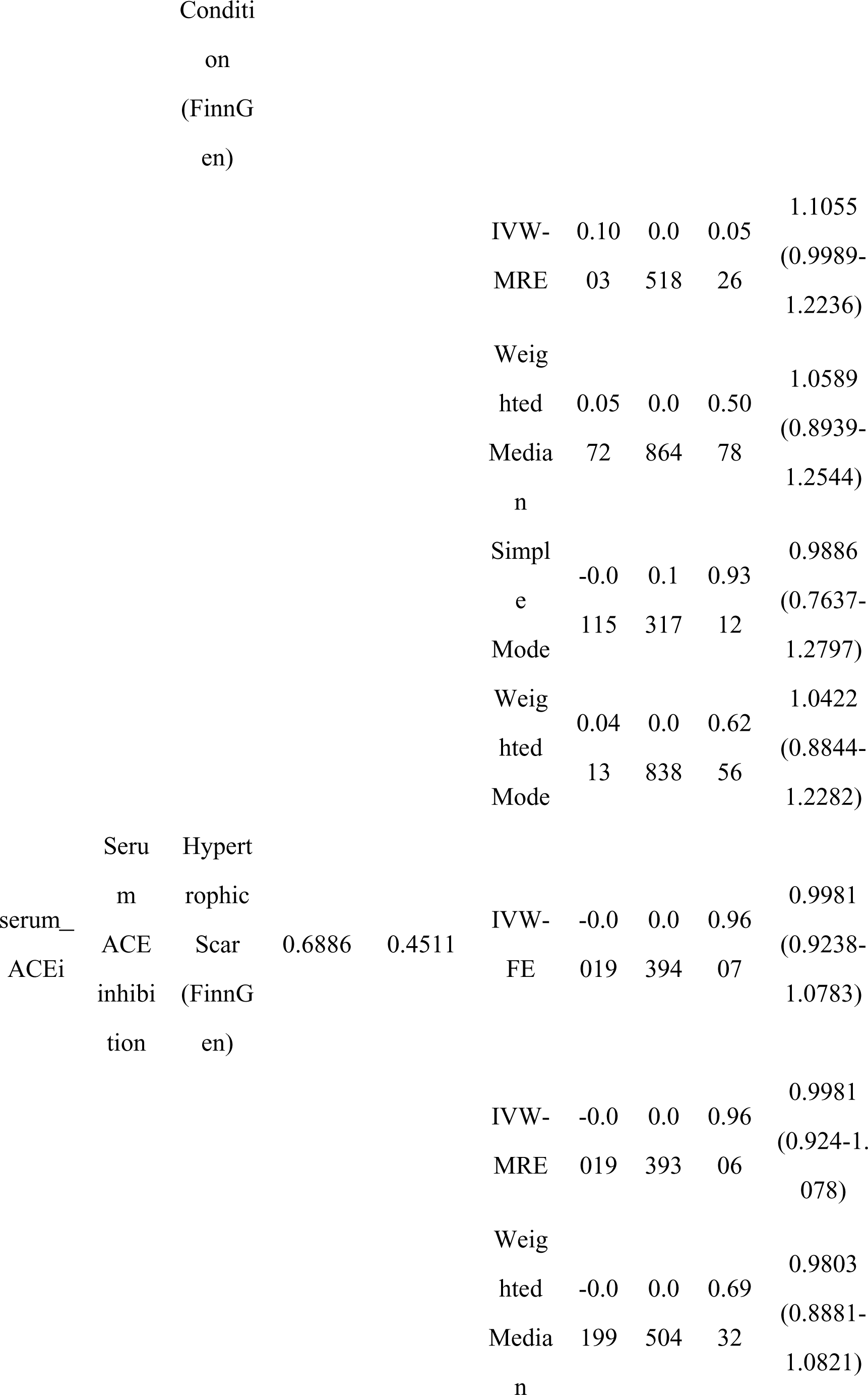
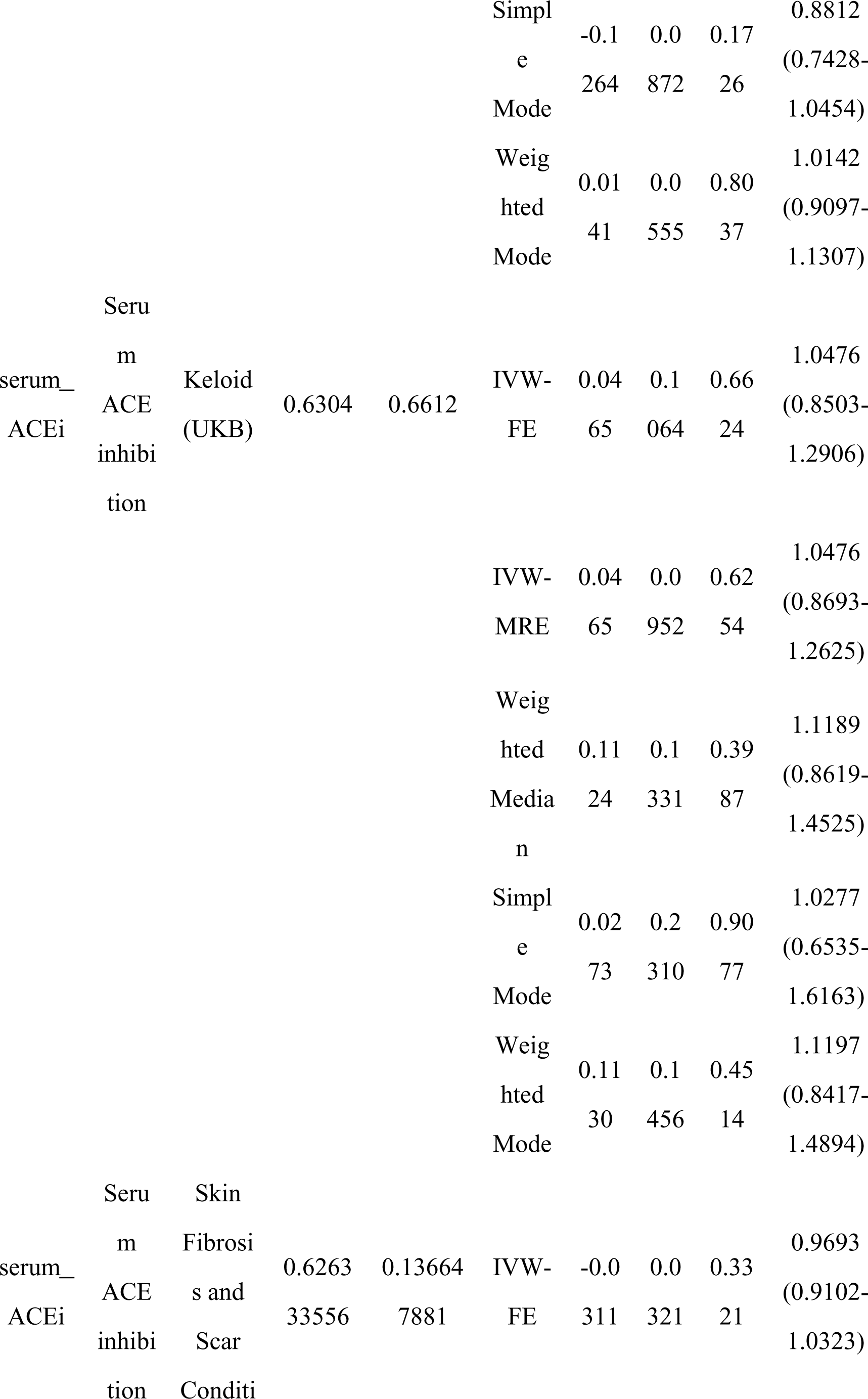
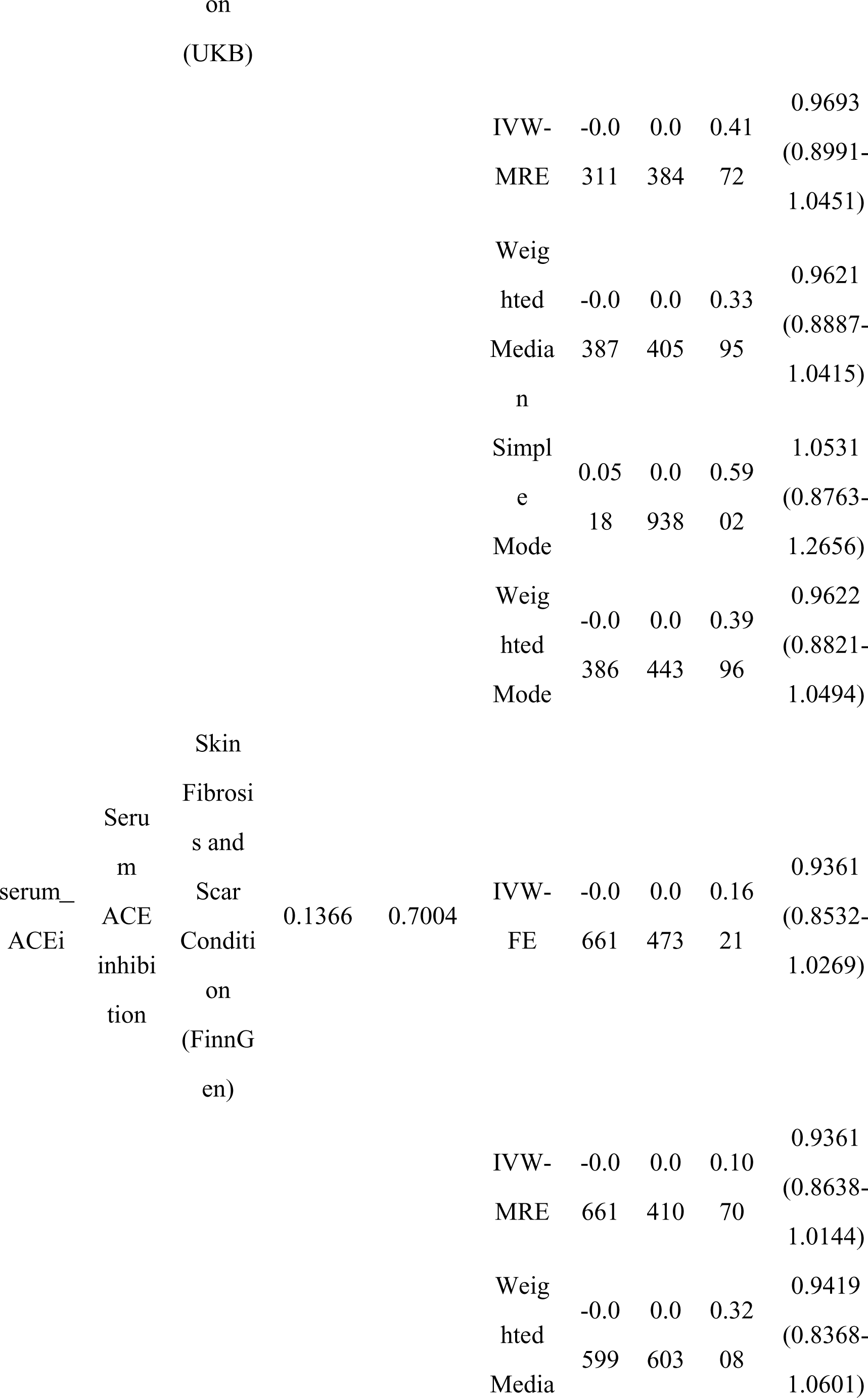
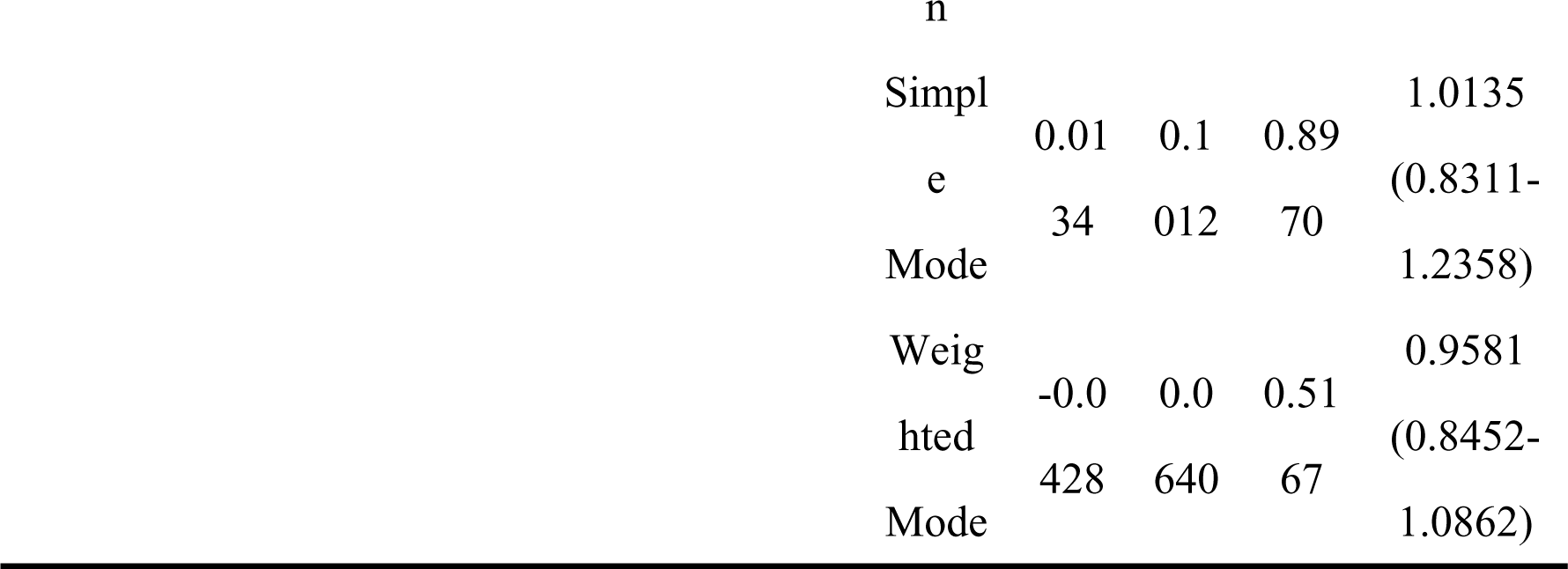
Serum ACE and Skin Fibrotic Diseases.

### Effect of blood pressure by ACE or ACE inhibition and skin fibrotic diseases

For both IVs of single SNP or multiple SNPs, results of Wald ratio and IVW-FE showed little causal association between the effect of ACE inhibition on decreasing blood pressure by on skin fibrotic diseases (hypertrophic scar, skin fibrosis and scar condition), all the p values were above 0.05 (shown in Table 4 and Figure 2). For MR analysis with IVs containing 2 SNPs, heterogeneity tests showed no significant heterogenetic bias existed in MR analysis (shown in Table 4).

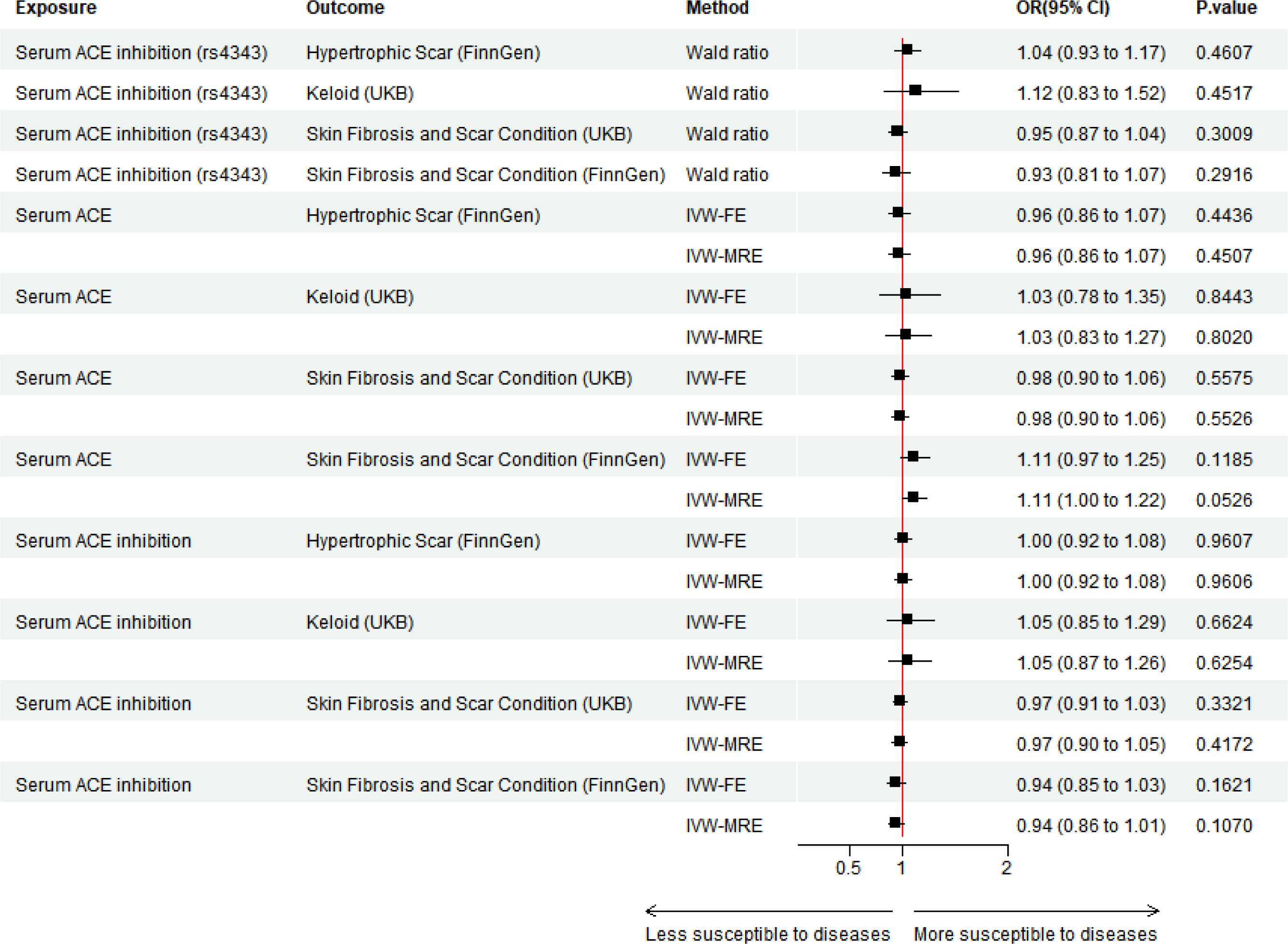

**Table 4.**
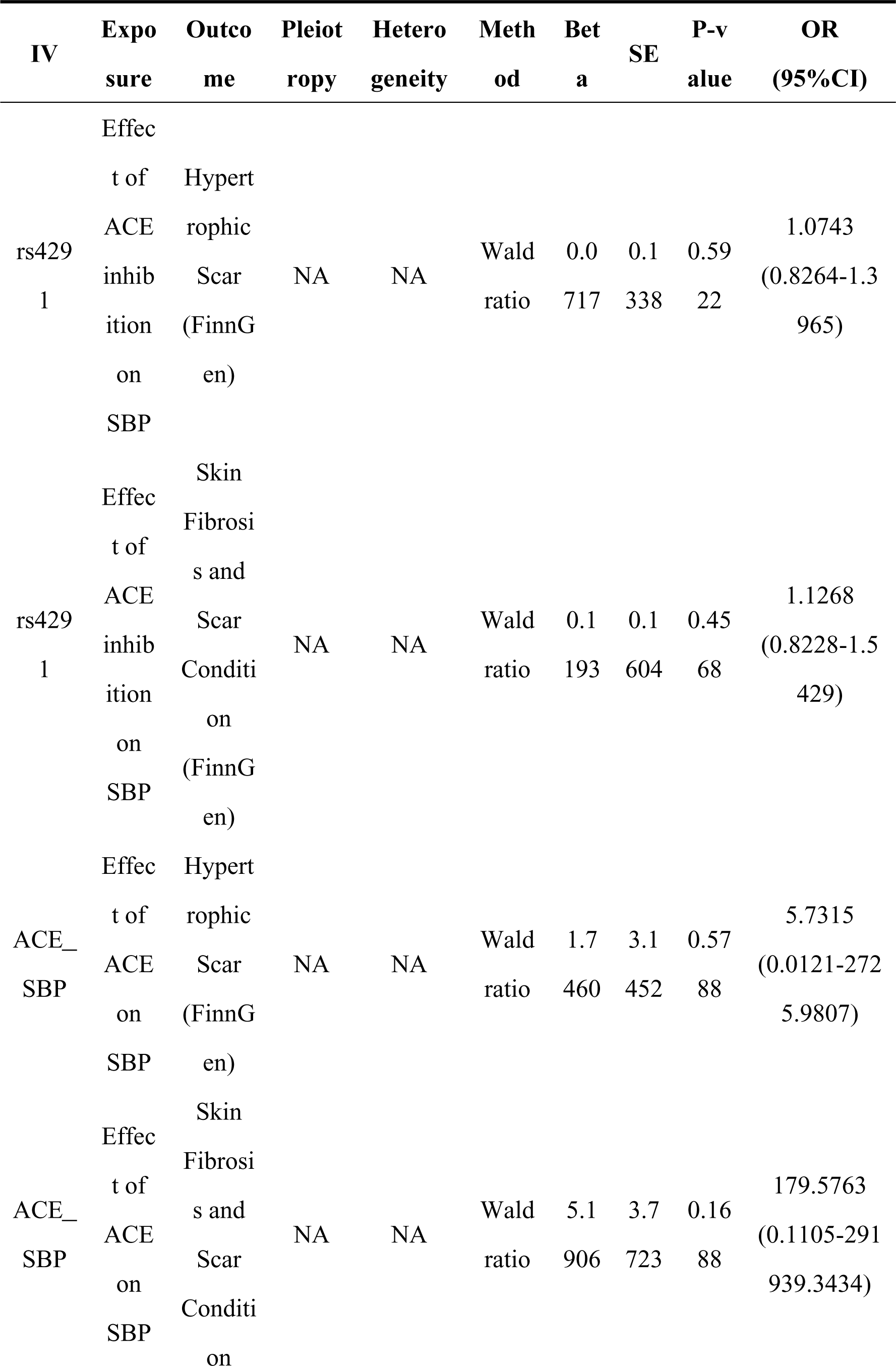
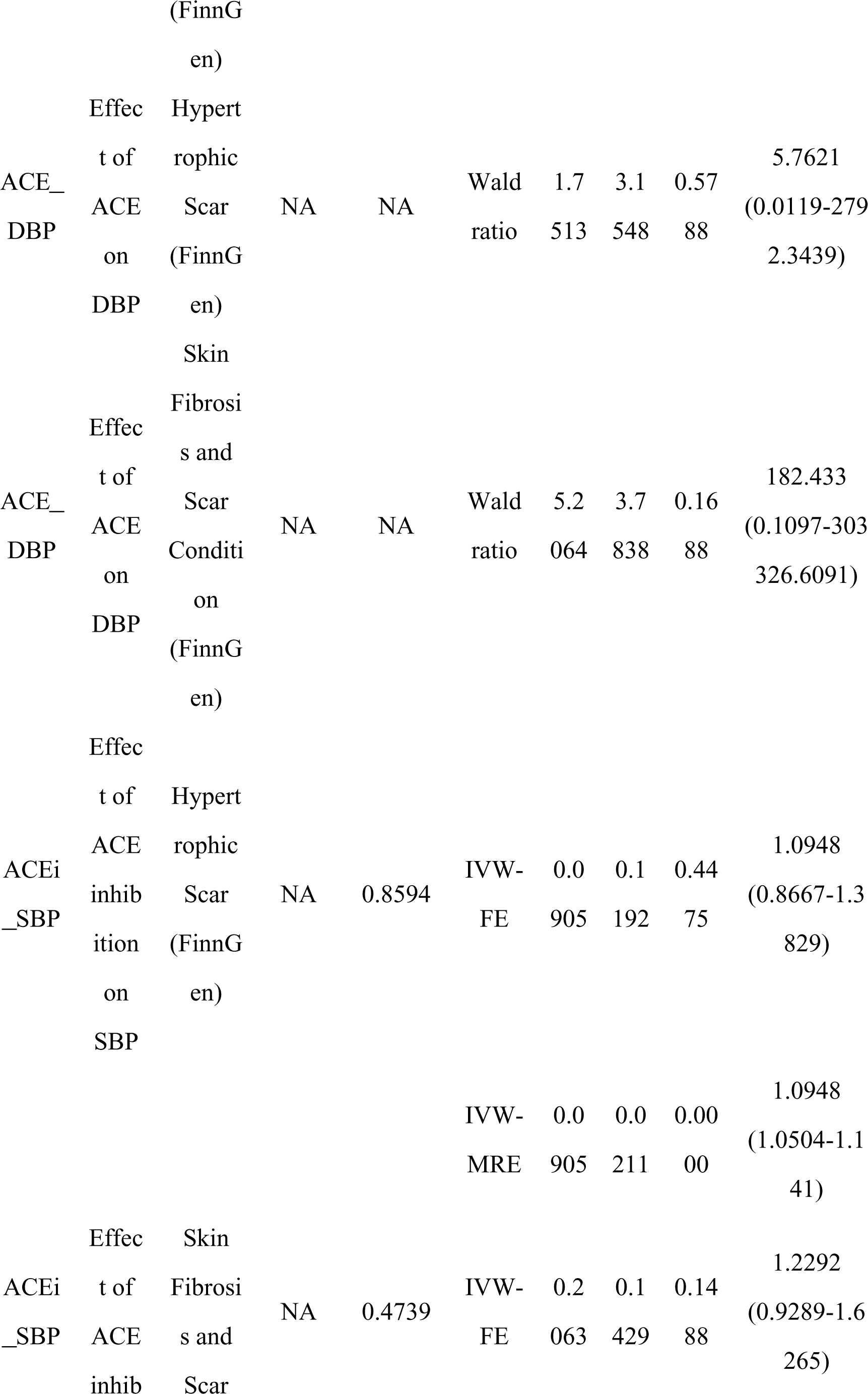
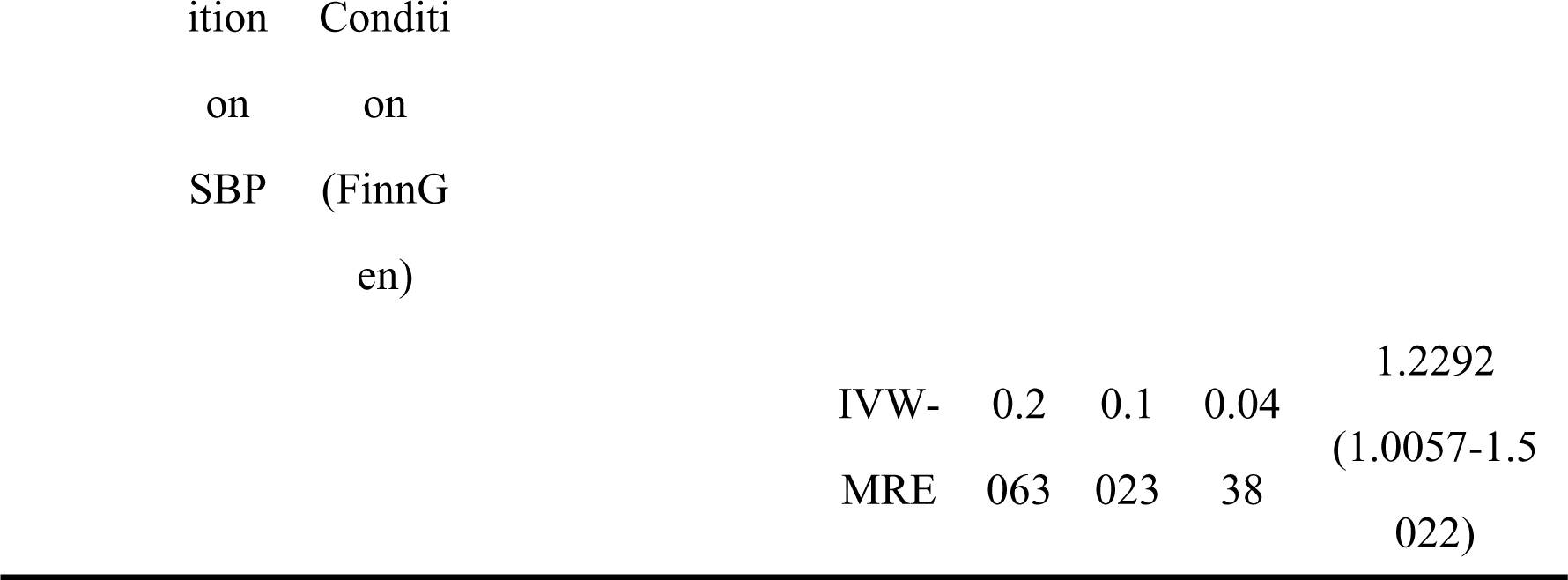
ACE Effect on Blood Pressure and Skin Fibrotic Diseases.

### Local ACE expression and skin fibrotic diseases

For skin tissue exposed to or shielded from sunshine exposure, 8 and 11 SNPs was extracted from Qtex datasets respectively to proxy local ACE expression, respectively. Notably, all the p values of SMR analysis were above 0.05 (shown in Table 5), which showed that little causal effect of ACE expression in local skin tissue on susceptibility of skin fibrotic diseases, irrespective of sunshine exposure. All the p values for HEIDI test were above 0.05 (shown in Table 5), which revealed that no significant heterogenetic bias existed.

**Table 5.**
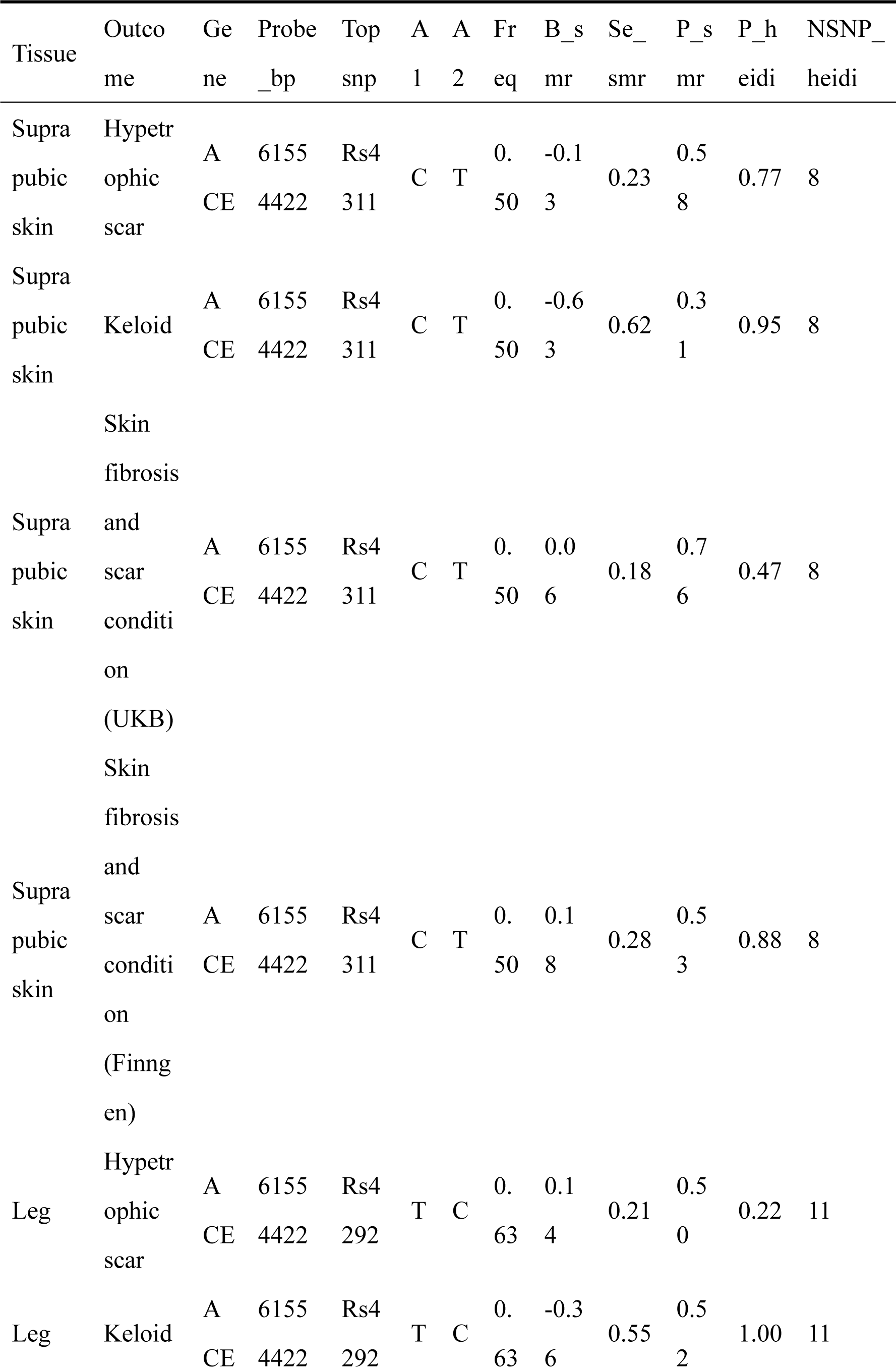
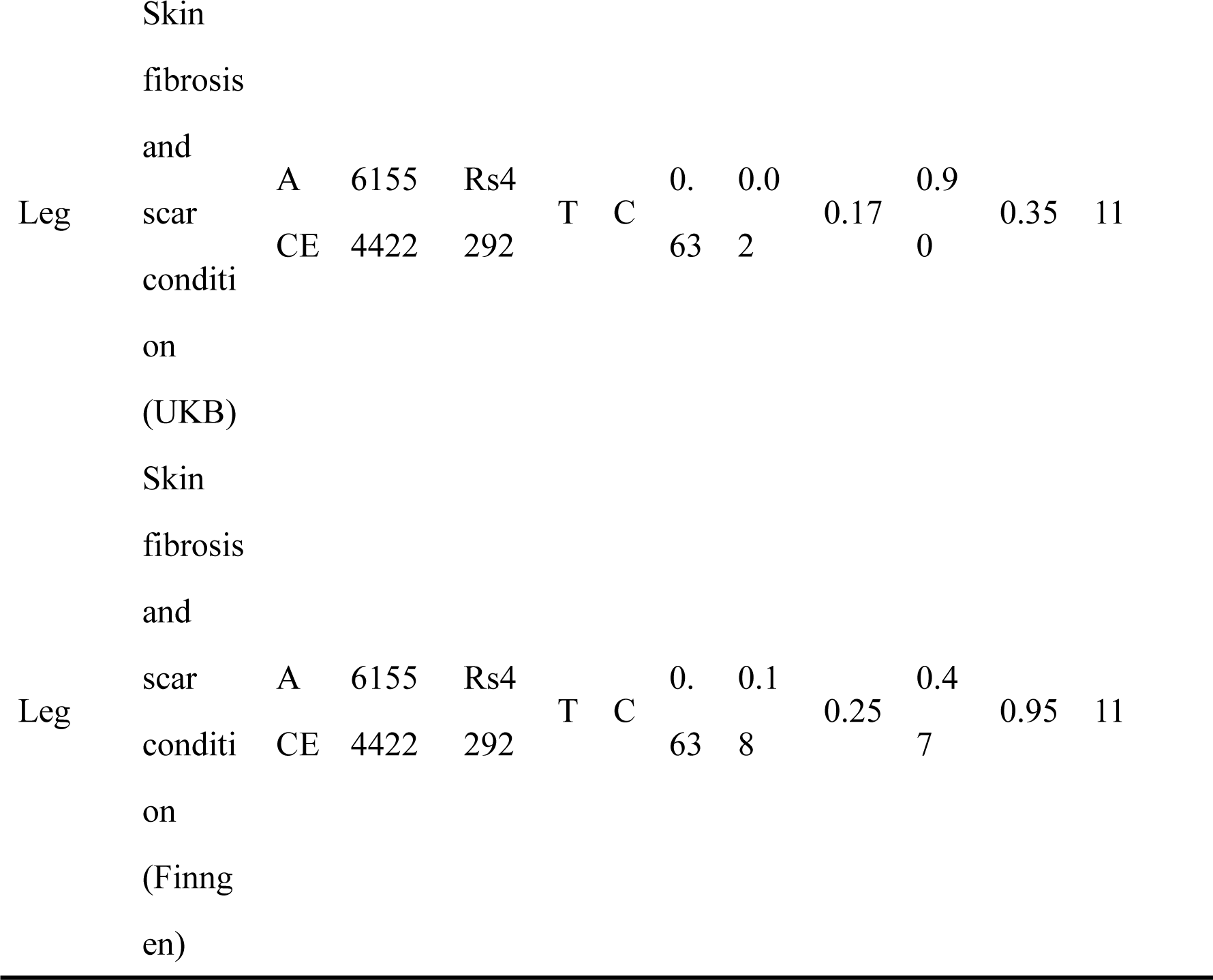
ACE Expression in Local Skin Tissue and Skin Fibrotic Diseases.

### Systolic and diastolic blood pressure and skin fibrotic diseases

Results of IVW-FE method showed significant negative causal effects of SBP on hypertrophic scar (OR = 0.69, 95% CI 0.54−0.89, p = 0.004). Similarly, results of MR Egger method, and IVW-MRE also demonstrated negative causal effect of SBP on hypertrophic scar, (MR Egger: OR = 0.480, 95% CI 0.244-0.947, p = 0.035; IVW-MRE: OR= 0.69, 95% CI 0.53-0.90, p = 0.007). These results collectively indicated that higher SBP level is causally associated with reduced risk of developing hypertrophic scar. However, IVW and other methods indicated little causal effect of DBP on hypertrophic scar. Additionally, no significant causal relationship was found between SBP or DBP and skin fibrosis and scar condition (shown in Table 6 and Figure 3). Sensitivity tests showed no obvious bias in pleiotropy and heterogenity.

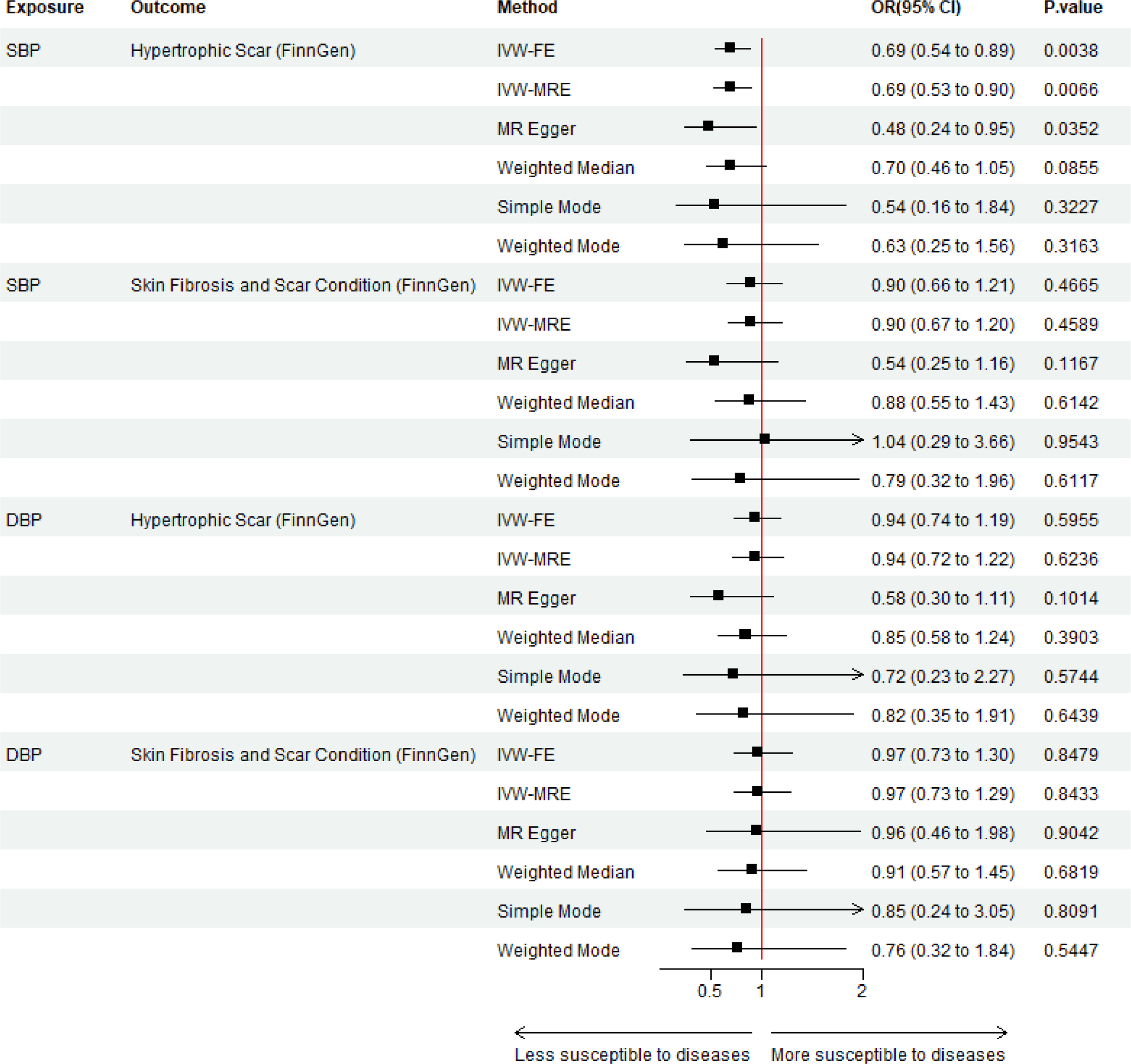

**Table 6.**
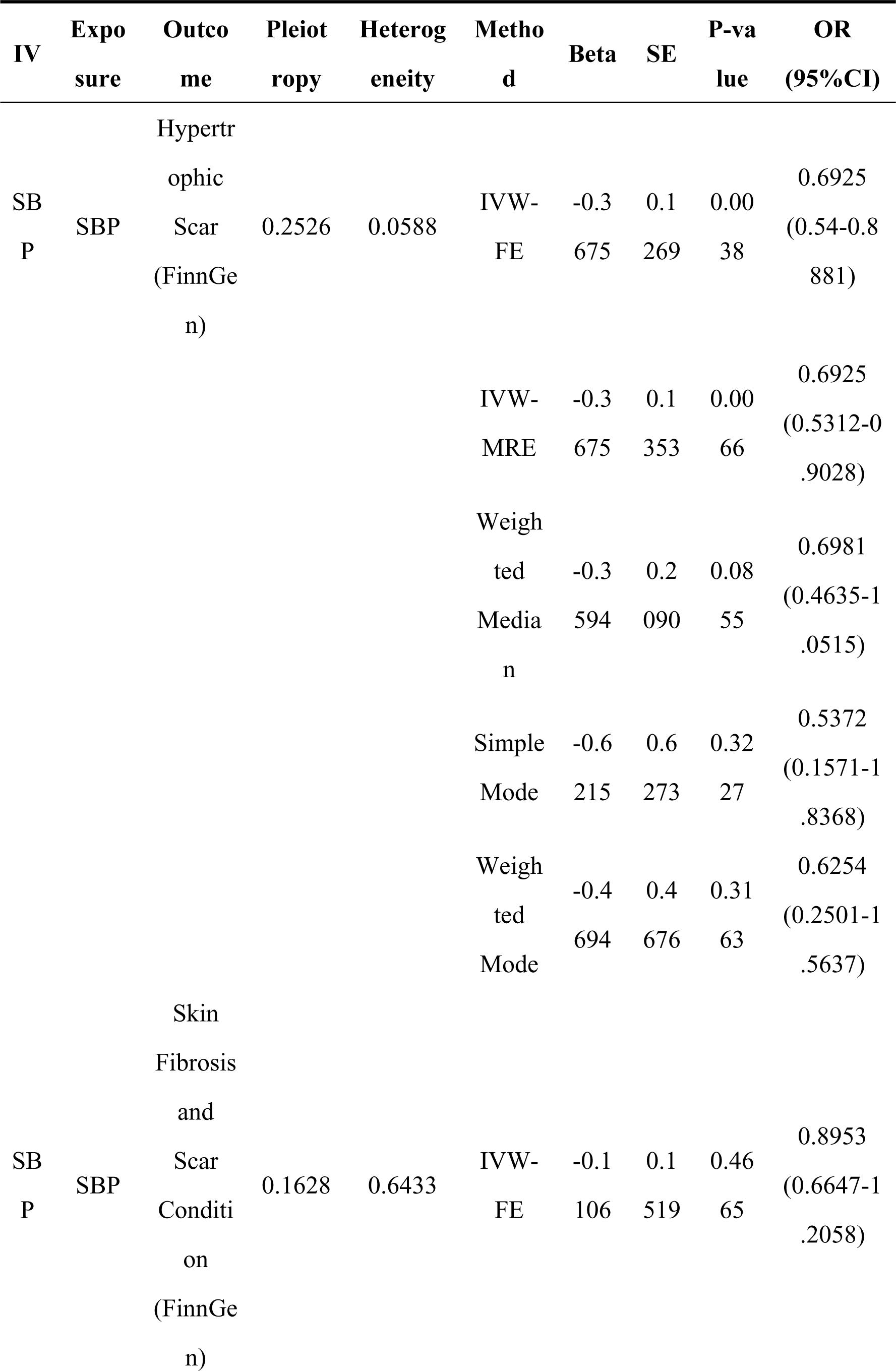
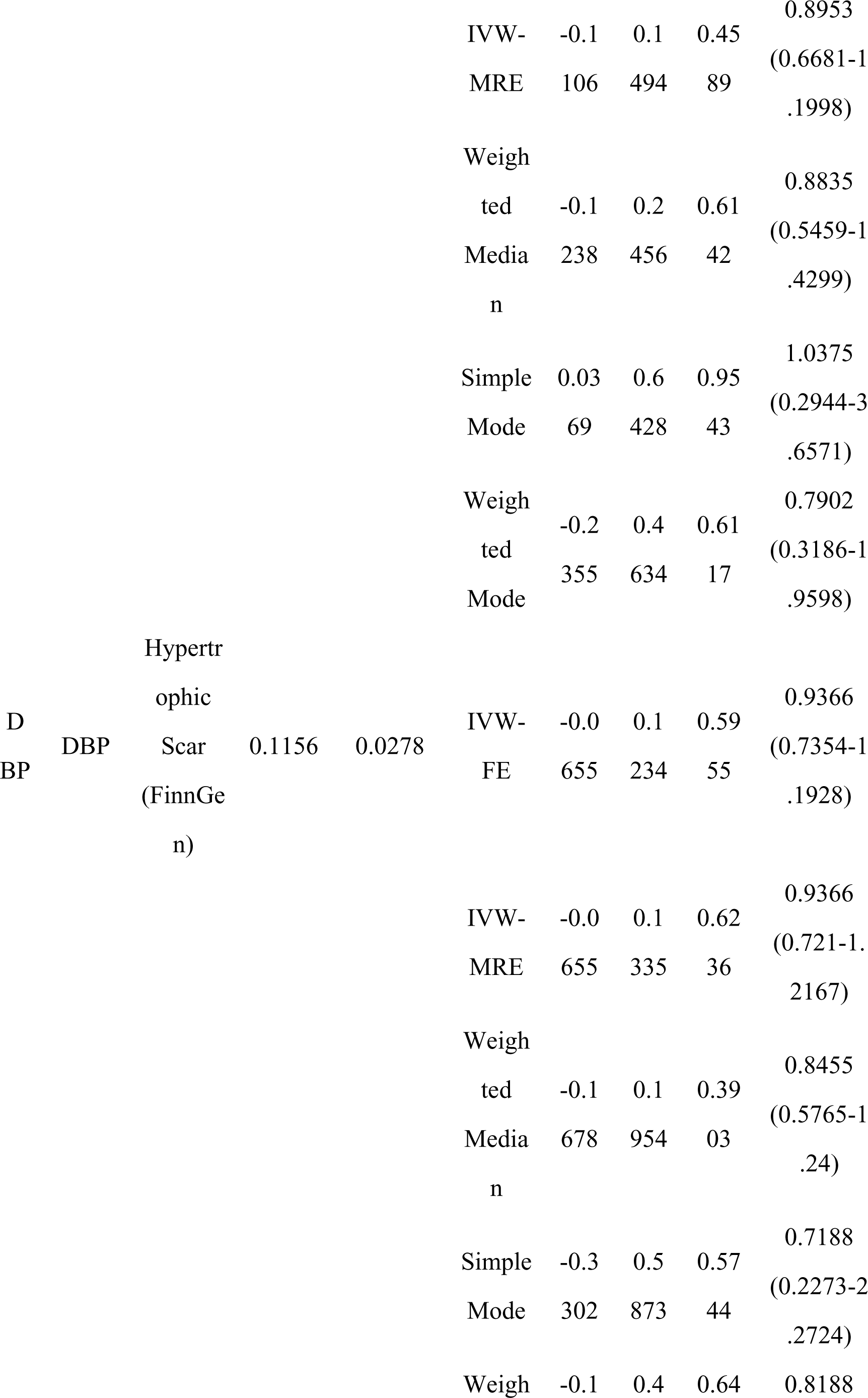
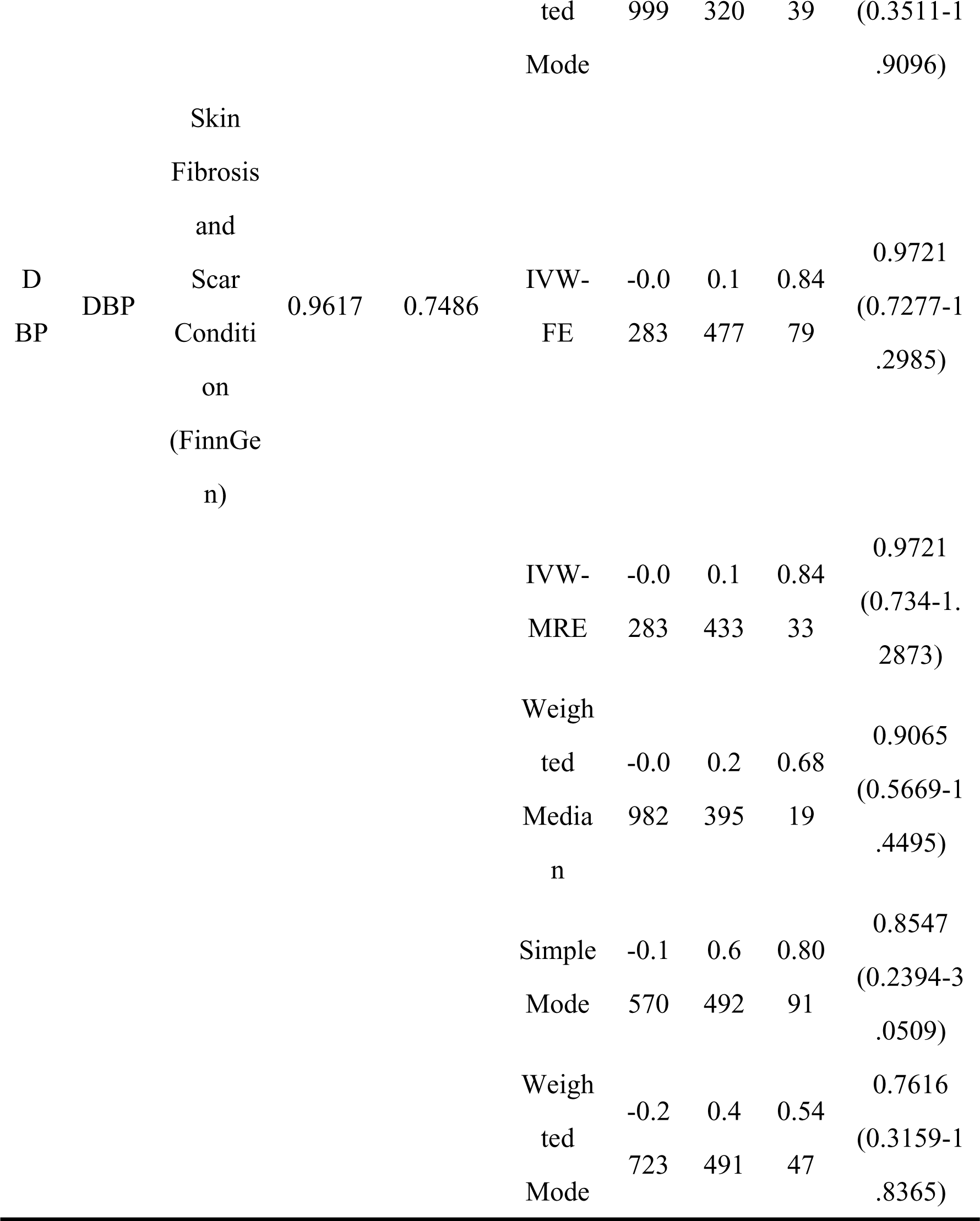
Blood Pressure and Skin Fibrotic Diseases.

### Effect of other antihypertension drugs and skin fibrotic diseases

Results of IVW-FE revealed obvious positive causal relationship between effect of BBs on decreasing SBP and skin fibrosis and scar condition from FinnGen (OR = 1.184, 95%CI 1.033-1.357, p = 0.015), which indicated that BBs caused an increase of risk of skin fibrosis. Heterogeneity and pleiotropy tests for both MR analysis showed little heterogeneity and horizontal pleiotropy (shown in Table 7 and Figure 4). However, results of IVW-MRE indicated little causal relationship between effect of BBs on decreasing SBP and hypertrophic scar, for which heterogenesis test revealed significant heterogeneity of this result (pQ = 0.008). And results of IVW-FE showed little causal relationship between effects of CCBs on decreasing SBP and hypertrophic scar or skin fibrosis and scar condition, either. Similarly, results of simple mode, weighted mode and weighted median methods for MR analysis showed similar conclusions of the little causal effect of CCBs on hypertrophic scar or skin fibrosis and scar condition. In heterogeneity and pleiotropy test, except MR between BBs and hypertrophic scar, no obvious pleiotropy and heterogeneity bias was observed. Wald ratio results found little causal relationship between blood decreasing effect of ARBs and skin fibrotic diseases (shown in Table 7 and Figure 4).

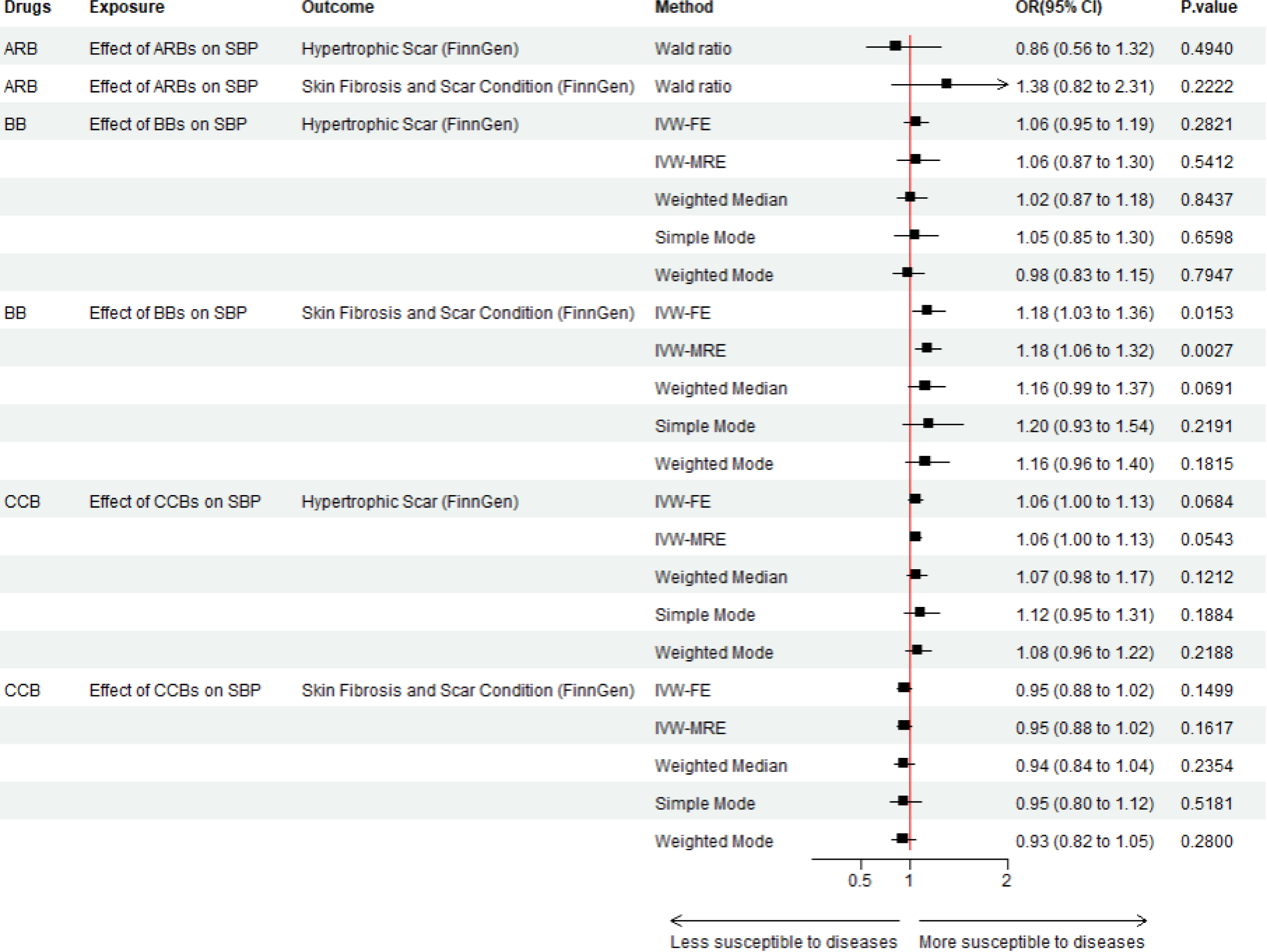

**Table 7.**
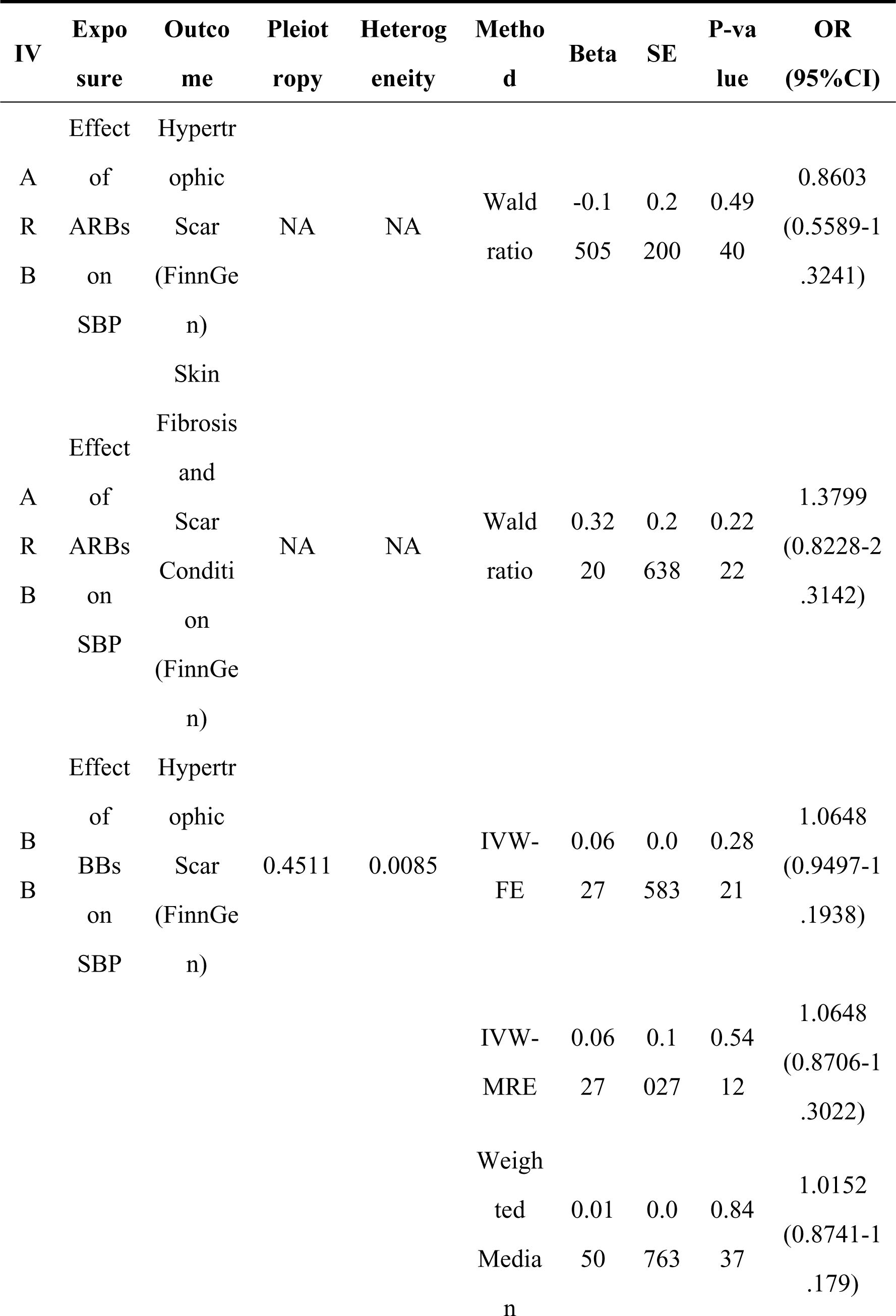
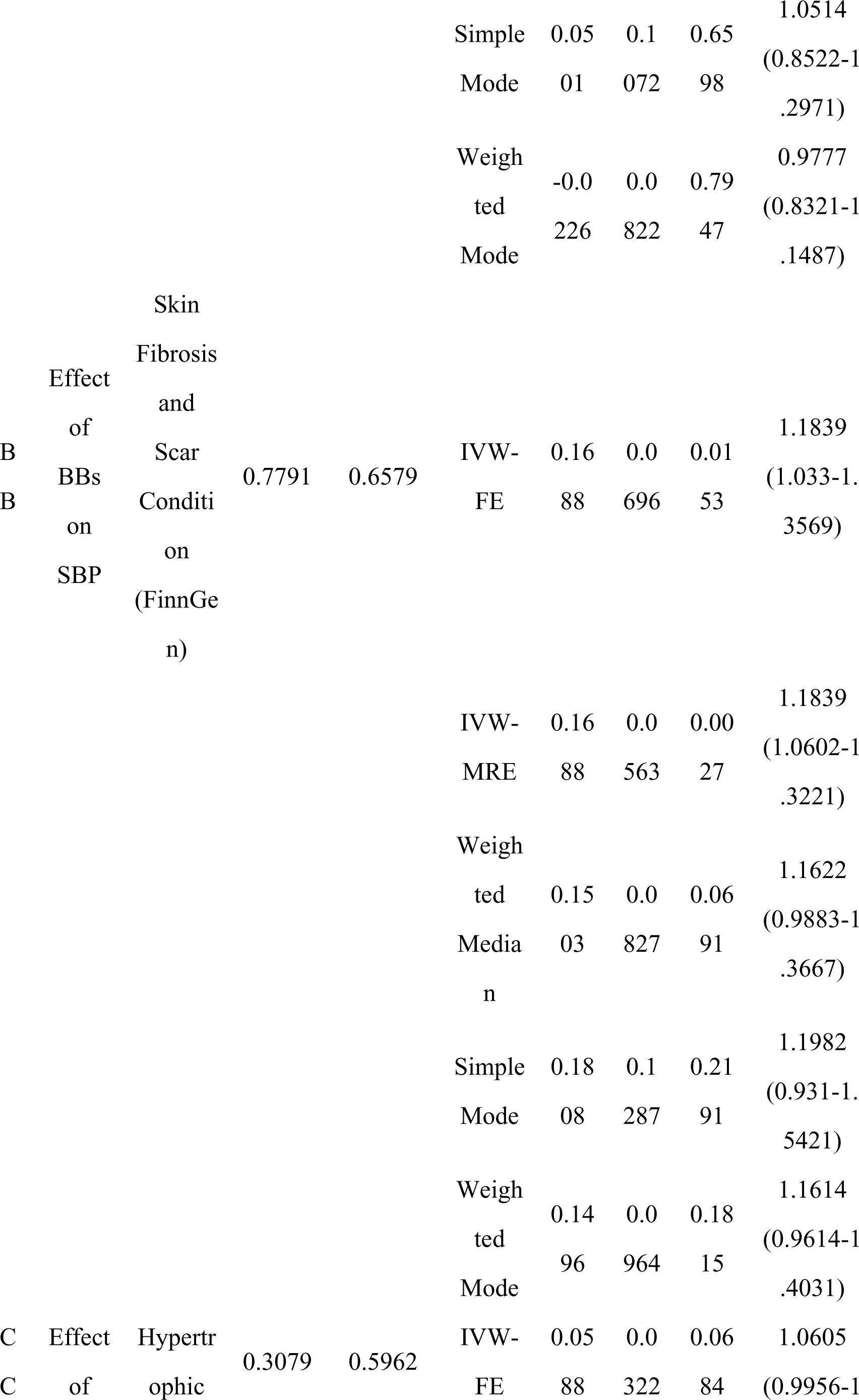
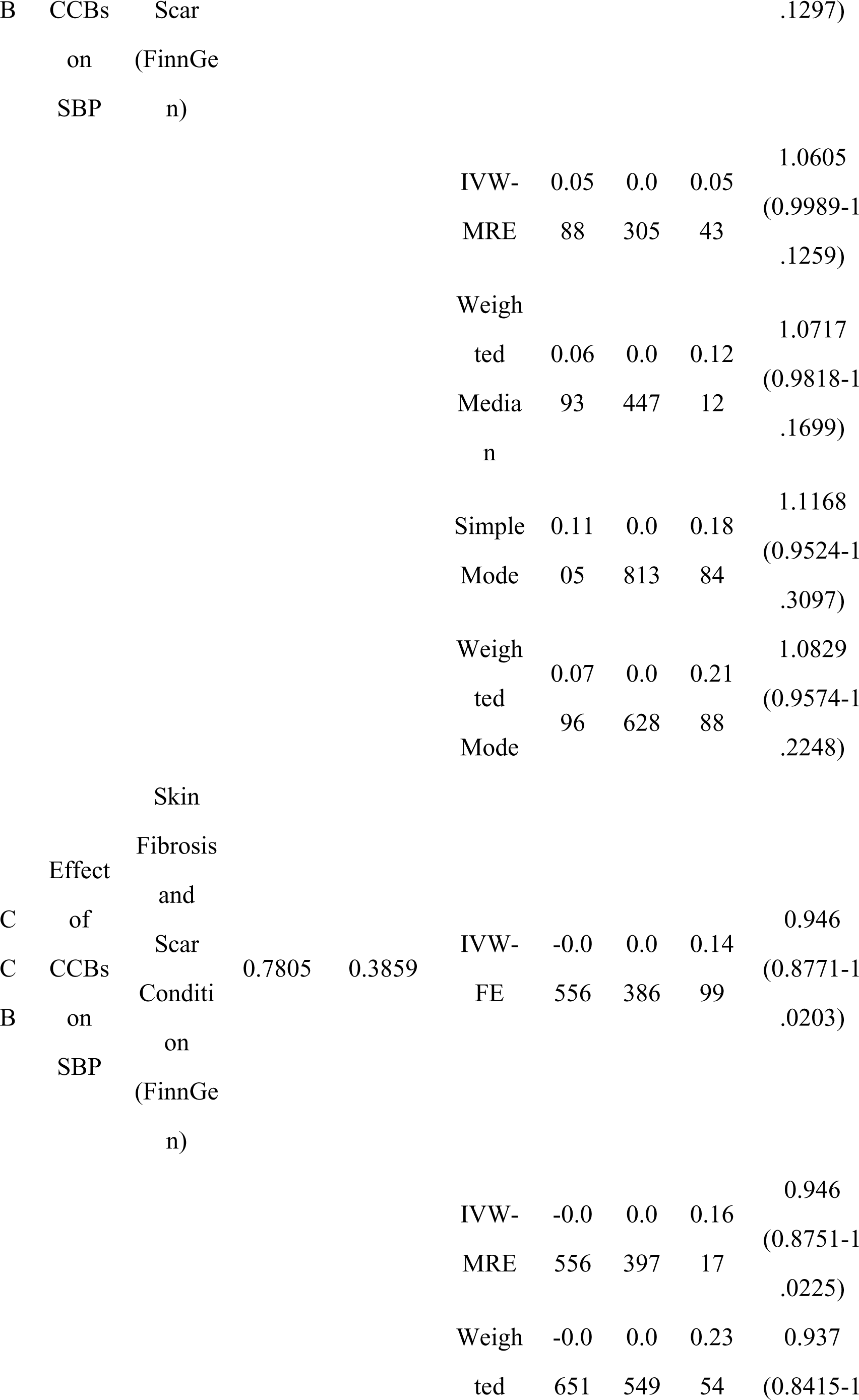
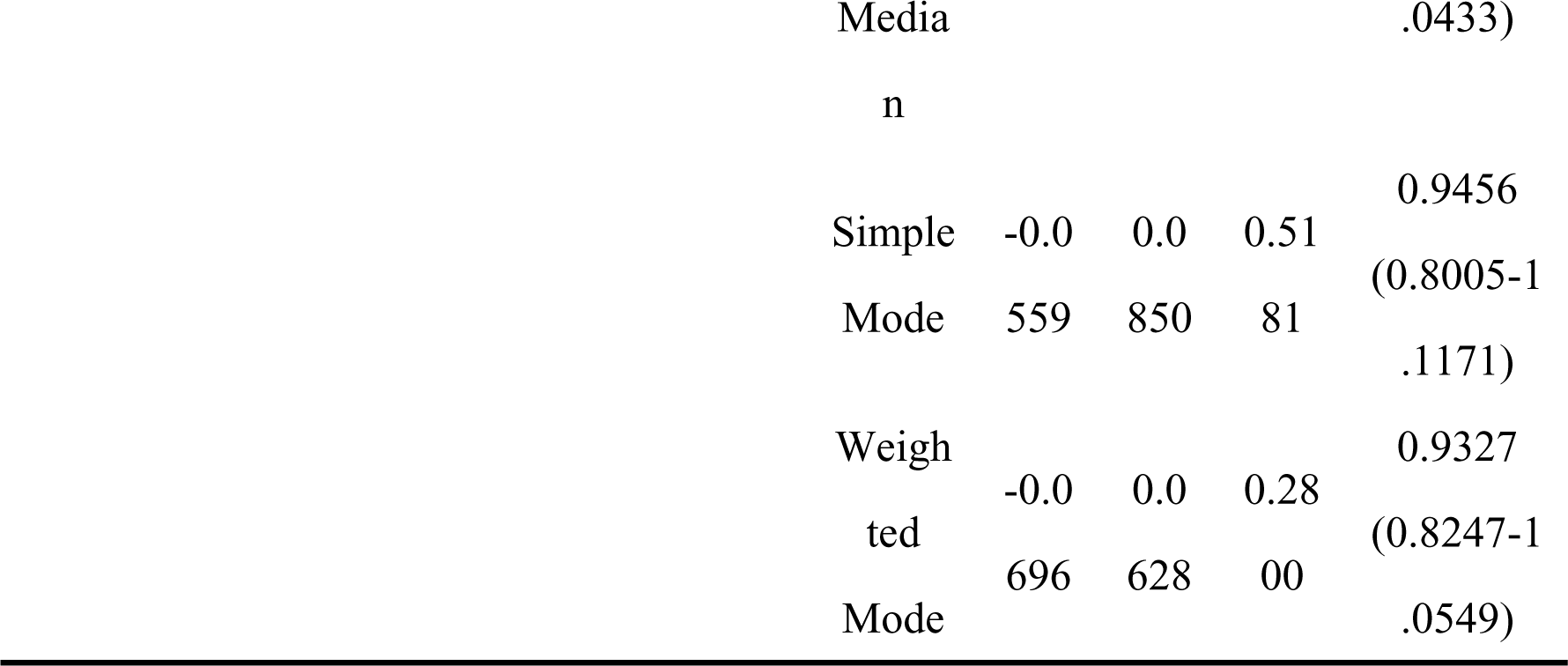
Other Antihypertension Drugs’ Effect on Blood Pressure and Skin Fibrotic Diseases.

## Discussion

Our study found little causal relationship of ACE inhibition on skin fibrotic diseases using two sample MR approach. None of serum ACE, effect of decreasing SBP/DBP by ACE inhibition or local expression of ACE gene in skin tissue was discovered significant with causal association with the occurrence of skin fibrosis and excessive scars including keloid and hypertrophic scar.

Nevertheless, a limited number of animal studies have suggested that ACE inhibitors can impede progression of fibrotic skin scars when administered orally or topically, indicating their potential as a therapeutic or preventive agent for such scars. ^11–13^ Additionally, a few case reports, ^43^ the aforementioned retrospective study ^14^ reported the potential of ACE inhibitor drugs (Enopril and Captopril) treatments for hypertrophic scar and keloid on patients. However, the isolated case reports and retrospective study did not exclude disrupting factors. Moreover, in the retrospective study, utilization of ACE inhibitor and ARB were mixed and the dosage were not unified. The aforementioned clinical trial conducted in Iran^15^ demonstrated the effectiveness of tropical ACE inhibitor (Enopril) on decreasing the size of hypertrophic scar. However, there was a lack of large scale clinical researches covered multiple populations which provided evidence for the efficacy of ACE inhibitors in prevention or treatment of skin fibrotic scars.

A potential explanation for the discrepancies in the findings lies in the diverse statistical criteria employed to define the outcomes. While animal studies and clinical trials assessed the outcomes based on scar area, our research utilized a GWAS dataset with a binary outcome measure. Our findings indicated that ACE inhibition does not decrease the incidence of skin fibrotic diseases, which is not conflictive with the conclusion that ACE inhibitors have potential to diminish the size of skin fibrotic scar. To the best of our knowledge, there are no quantitative GWAS datasets available with phenotype specially focused on the size of fibrotic scar.

Furthermore, the regulation of fibrotic proliferation in tissue by ACE is not a unidirectional process. The products of ACE, angiotensin II, exhibits dual effects on fibrosis. When angiotensin II bind to AT1 receptor, its downstream effect is to promote fibrosis by promoting inflammation reactions such as TGF-β synthesis. When angiotensin II binding to AT2 receptor, it can counteract inflammation reactions and suppress reactions like TGF-β synthesis. Additionally, other downstream products of angiotensin II, such as angiotensin III and angiotensin IV, also have the potential to inhibit fibrosis. ^44, 45^

It is noted that the expression of AT1 receptors in skin is comparatively low. RNA seq from normal tissue of different organs indicated that reads per kilobase per million mapped reads (RPKM) of AT1 receptor in skin is 0.906 ± 0.285, significantly lower than that in other tissues such as liver (21.967 ± 2.326), kidney (5.741 ± 1.363) and heart (2.423 ± 1.416). ^46^ These results prompt that the regulation effect by ACE in skin might be less pronounced than that in other tissue or organs.

Compared to previous studies, our research had several advantages: 1) Our study provided an exploration of the association between ACE and skin fibrotic diseases with a larger sample scale in European population. 2) Our study design had advantages of eliminating confounding factors as much as possible by pleiotropy test and LD diminishing. 3) Our study investigated the causal relationship beyond merely association relationship by the method of MR analysis.

Our research also existed some limitations. The conclusion that ACE inhibition has little causal effect on skin fibrosis was not fully confirmed due to several limitations: 1) All the MR analysis were conducted within a single population of European descent. To validate the effect of ACE inhibition on skin fibrotic diseases in other population, GWAS researches from corresponding population were required. 2) The statistical power of MR was insufficient, necessitating GWAS studies of skin fibrotic diseases with larger sample sizes. To the best of our knowledge, exposures for effect of BBs, CCBs and ARBs lacked data of pQTLs of corresponding protein targets, and the analysis primarily included the effects of decreasing blood pressure. Consequently, the direct effects of those protein targets require further research to determine whether significant causal relationship exists between their effects and skin fibrotic diseases.

## Conclusion

Our findings suggest that ACE inhibitors do not have a direct causal effect on the risk of skin fibrotic diseases, including hypertrophic scars and keloids. This challenges the potential of ACE inhibitors as a therapeutic option for preventing or treating these conditions. Further researches including large scale clinical trials and mechanisms researches are needed to explore the complex interplay between RAS system and skin fibrosis and the potential appications of RAS system on skin fibrotic diseases.

## Fund

Beijing Tsinghua Changgung Hospital Fund (Grant No. 12023C1004)

## Ethical Approval

Not Applicable.

## Ethnic Statement

Not Applicable.

## Data Availability Statement

Sources of full summary GWAS datasets were listed in Supplementary File 2. No new data was generated from recurrent study.

## Author Contributions

Conceptualization: The study was initially conceptualized by YW, ZW and JT. They developed the research idea and designed the overall framework of the study.

Methodology: Methodology was developed by ZW, YJ and ZL. They were responsible for designing and implementing the experimental protocols.

Investigation: Investigation and data collection were primarily conducted by YW, ZW and JT. They carried out the experiments and gathered the necessary data.

Data Analysis: Data analysis was performed by YW and ZW. They processed the raw data and conducted the statistical analyses.

Writing - Original Draft Preparation: The original draft of the manuscript was written by YW and ZW. They prepared the initial version of the paper, including the introduction, methods, results, and discussion sections.

Writing - Review & Editing: Review and editing of the manuscript were conducted by YJ, ZL, MY and JT. They reviewed the draft for important intellectual content and made revisions.

Funding Acquisition: Funding for the study was secured by MY and JT. They applied for and obtained the necessary grants to support the research.

Supervision: Supervision of the entire project was provided by MY and JT. They oversaw the project progress and provided guidance throughout the research.

Project Administration: Project administration, including coordination of research activities and maintaining timelines, was managed by MY and JT.

